# The anthropogenic fingerprint on emerging infectious diseases

**DOI:** 10.1101/2024.05.22.24307684

**Authors:** Rory Gibb, Sadie J. Ryan, David Pigott, Maria del Pilar Fernandez, Renata L. Muylaert, Gregory F. Albery, Daniel J. Becker, Jason K. Blackburn, Hernan Caceres-Escobar, Michael Celone, Evan A. Eskew, Hannah K. Frank, Barbara A. Han, Erin N. Hulland, Kate E. Jones, Rebecca Katz, Adam Kucharski, Direk Limmathurotsakul, Catherine A. Lippi, Joshua Longbottom, Juan Fernando Martinez, Jane P. Messina, Elaine O. Nsoesie, David W. Redding, Daniel Romero-Alvarez, Boris V. Schmid, Stephanie N. Seifert, Anabel Sinchi, Christopher H. Trisos, Michelle Wille, Colin J. Carlson

## Abstract

Emerging infectious diseases are increasingly understood as a hallmark of the Anthropocene^1–3^. Most experts agree that anthropogenic ecosystem change and high-risk contact among people, livestock, and wildlife have contributed to the recent emergence of new zoonotic, vector-borne, and environmentally-transmitted pathogens^1,4–6^. However, the extent to which these factors also structure landscapes of human infection and outbreak risk is not well understood, beyond certain well-studied disease systems^7–9^. Here, we consolidate 58,319 unique records of outbreak events for 32 emerging infectious diseases worldwide, and systematically test the influence of 16 hypothesized social and environmental drivers on the geography of outbreak risk, while adjusting for multiple detection, reporting, and research biases. Across diseases, outbreak risks are widely associated with mosaic landscapes where people live alongside forests and fragmented ecosystems, and are commonly exacerbated by long-term decreases in precipitation. The combined effects of these drivers are particularly strong for vector-borne diseases (e.g., Lyme disease and dengue fever), underscoring that policy strategies to manage these emerging risks will need to address land use and climate change^10–12^. In contrast, we find little evidence that spillovers of directly-transmitted zoonotic diseases (e.g., Ebola virus disease and mpox) are consistently associated with these factors, or with other anthropogenic drivers such as deforestation and agricultural intensification^13^. Most importantly, we find that observed spatial outbreak intensity is primarily an artefact of the geography of healthcare access, indicating that existing disease surveillance systems remain insufficient for comprehensive monitoring and response: across diseases, outbreak reporting declined by a median of 32% (range 1.2%-96.7%) for each additional hour’s travel time from the nearest health facility. Our findings underscore that disease emergence is a multicausal feature of social-ecological systems, and that no one-size-fits-all global strategy can prevent epidemics and pandemics. Instead, ecosystem-based interventions should follow regional priorities and system-specific evidence, and be paired with investment in One Health surveillance and health system strengthening.

## Introduction

In the last few decades, emerging infectious diseases transmitted by wildlife (zoonoses; e.g. COVID-19, Ebola virus disease, influenza, and mpox) or arthropod vectors (e.g. dengue fever, Lyme disease, and Zika virus disease) have had catastrophic social, economic and ecological impacts. This trend runs counter to overall improvements in population health, and is widely believed to be the result of an ongoing state shift in the biosphere^14,15^, where human-driven environmental change has both increased animal susceptibility to infection, and created more opportunities for animal-to-human transmission (zoonotic spillover^2^), leading to more outbreaks of both familiar and novel pathogens. The rising tide of emerging infectious diseases has brought global attention to ecological and social interventions that could mitigate the upstream drivers of disease emergence^13,16^. Recently, most attention has focused on curbing wildlife trade or deforestation^17–20^, but other interventions could include greenhouse gas emissions reduction to limit climate change, human and livestock vaccination, improved access to point-of-care diagnostics and clinical care, the development of “One Health” disease surveillance systems and workforces, and stricter biosafety and biosecurity practices^17,21,22^.

Although these interventions are grounded in public health and ecological first principles, there is limited scientific consensus on their potential benefits and relative priority, in large part because of insufficient evidence about the universality of many drivers of disease transmission and emergence. A growing number of literature syntheses and meta-analyses have found evidence of predictable anthropogenic impacts on disease dynamics in wildlife hosts of emerging infectious diseases^23,24^, as well as vertebrate host^25–27^ and arthropod vector^27–29^ community composition. In general, these studies suggest that habitat fragmentation and disturbance, biodiversity loss, and agriculture tend to increase wildlife disease prevalence^30^, but the net impacts of urbanization, deforestation, and climate change may be more unpredictable^31–34^. This reflects a mix of scientific evidence gaps and true heterogeneity across systems, driven by differences in pathogen life cycles, host and vector ecology, and the intensity and types of anthropogenic impacts. As a result, the downstream relationship between human disease risk and climate change, biodiversity loss, or land use may also be idiosyncratic across diseases and regions^35^. These relationships are further complicated by exposure processes: human-wildlife contact patterns and social vulnerability to outbreaks both vary across landscapes and populations, and neither are usually captured in wildlife-focused studies.

A few influential studies have directly examined the ecological correlates of human disease emergence, based on the location where and circumstances under which around 300 emerging pathogens were first scientifically identified (“emergence events”)^1,4–6^. These studies have found that land use change, agricultural expansion, biodiversity hotspots and global travel are widely associated with observed geographic hotspots of disease emergence. However, the historical circumstances of the first confirmed outbreak may not be representative of the social and ecological factors that determine wider landscapes of infection risk. By design, data on emergence events are biased towards better-resourced settings where new diseases are more likely to be detected and described, rather than the rural, poor, and marginalized populations in lower-resource settings that experience an endemic burden of zoonotic and vector-borne diseases^36^ (including many infections typically framed as “emerging”^37–39^). Assigning outbreak drivers based on expert opinion^5,6^ is also prone to confirmation bias^40^, and difficult to generalize to wider patterns of risk. The growing availability of fine-scale, comprehensive georeferenced outbreak datasets for many of these diseases – compiled from disease surveillance reports, scientific and gray literature^41^ -- provides the opportunity for a more systematic, global, data-driven assessment of emerging disease drivers.

In this study, we harmonized spatially-explicit human outbreak data sources for 32 emerging infectious diseases with available data (Figure 1, Extended Data Table 1, Supp. Table 1), including bat viruses with epidemic and pandemic potential (e.g. filo-, henipa-, and coronaviruses), rodent-borne pathogens (e.g. hanta- and arenaviruses, plague, and mpox), mosquito-borne arboviruses (including flavi-, alpha-, and orthobunyaviruses: e.g., chikungunya, dengue fever, and Rift Valley fever), and other neglected zoonotic, vector-borne, and environmentally-transmitted infections (e.g. melioidosis, Crimean-Congo hemorrhagic fever, and *Plasmodium knowlesi* zoonotic malaria). Because our goal was to understand drivers of outbreak risk, rather than the distinct factors that predispose outbreaks to become epidemics or pandemics, we only examined records associated with some level of environmental influence: we included all available records associated with vector-borne or environmental exposure, but for diseases with substantial onward human-to-human transmission chains (e.g. Ebola virus disease, mpox), only index cases with a probable zoonotic origin were included (Methods). Datasets were mainly collated from published scientific datasets, as well as national notifiable disease surveillance system data from the United States, Brazil, and Argentina. The complete dataset includes 58,319 unique outbreak events across 169 countries (Figure 1a; Methods) spanning 1910 to 2022 (but primarily post-2000; Figure 1b), with an outbreak event defined as ≥1 confirmed case at a given georeferenced location in a given year (either point or administrative polygon; Methods). Because of several source datasets’ focus on comprehensive coverage for certain diseases^42–54^, our harmonized database has widespread coverage in the Americas, sub-Saharan Africa and South and Southeast Asia, although records are still sparse in North Africa and above the 50° N latitude line.

**Figure 1.**
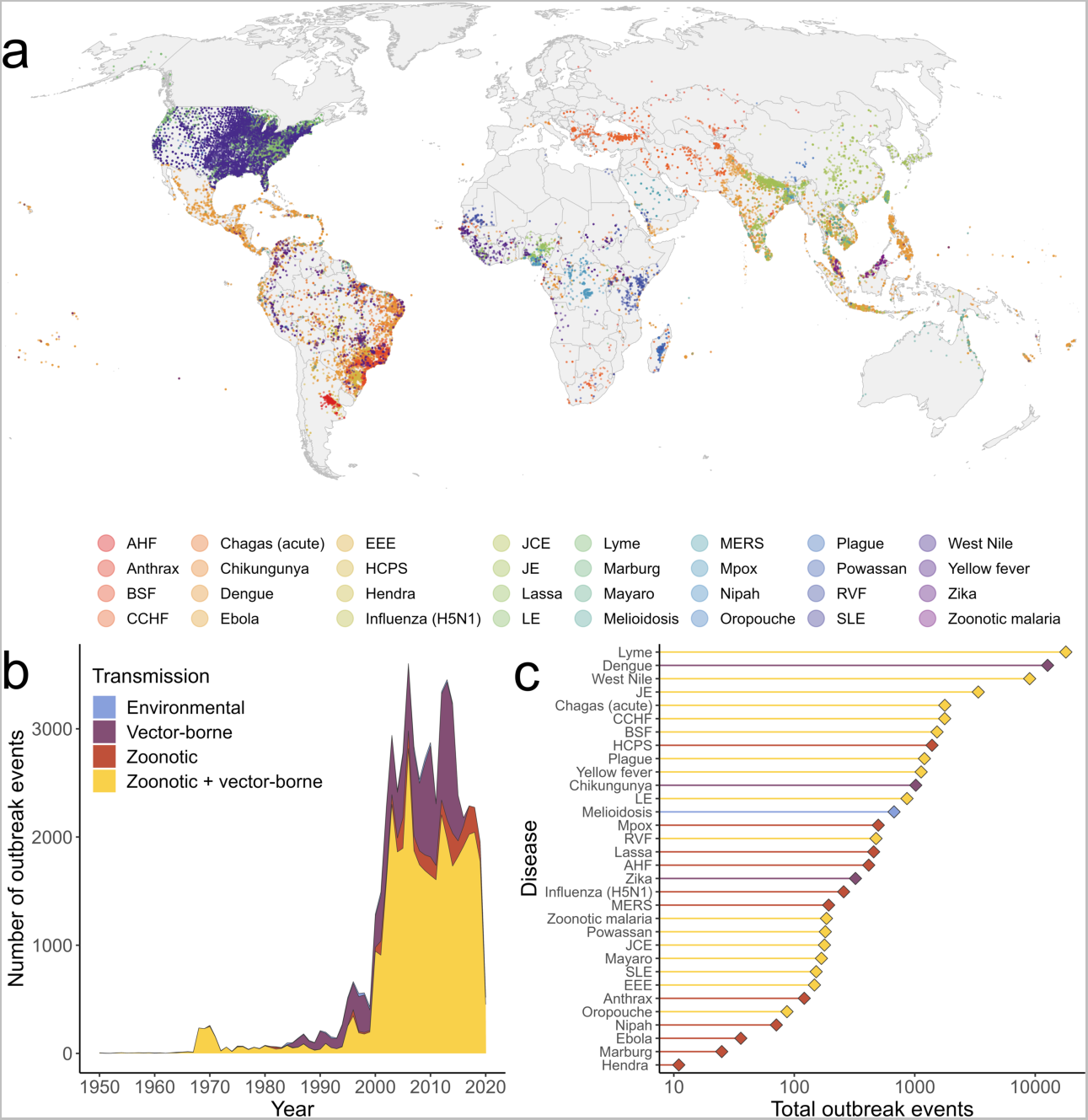
A global compendium of outbreak events for 32 human emerging infectious diseases. Records include a mix of georeferenced human disease occurrence or outbreak data and case incidence data from national surveillance systems (Extended Data Table 1) (A). Each point represents an outbreak event (at least 1 confirmed case per named locality per year) for diseases whose predominant human infection routes are broadly classified as either zoonotic (e.g. Ebola virus disease, Lassa fever), zoonotic and vector-borne (e.g. West Nile fever, yellow fever), vector-borne and mainly maintained in human hosts (e.g. dengue fever, chikungunya, and Zika virus disease), or transmitted through the environment (melioidosis) (B-C). Data were predominantly from post-2002 across all transmission types (B; data shown from 1950 onward), with the most data available for well-monitored widespread diseases (e.g. Lyme disease, dengue fever, West Nile fever) and the least for emerging bat-borne infections (filo- and henipaviruses) (C). Disease name abbreviations: AHF - Argentine hemorrhagic fever; BSF - Brazilian spotted fever; CCHF - Crimean-Congo hemorrhagic fever; EEE - Eastern equine encephalitis; HCPS - Hantavirus cardiopulmonary syndrome; JCE - Jamestown Canyon encephalitis; JE - Japanese encephalitis; LE - LaCrosse encephalitis; MERS - Middle East Respiratory Syndrome; RVF - Rift Valley fever; SLE - St. Louis encephalitis. For shorthand, we also omit “disease,” “virus disease,” and “fever” from disease names as appropriate; see Extended Data Table 1.

Using this extensive dataset, we developed a standardized framework for inferring the socio-environmental drivers of spatial outbreak intensity (Methods, Extended Data Figure 1). We use a modified case-control design^55,56^, which compares contemporary (post-1980) outbreak localities to population-weighted background locations (Methods, Extended Data Figure 2). At each location we extracted a set of 16 covariates from gridded geospatial datasets (Extended Data Table 2), which fall under five broad categories: *detection processes* (motorized travel time to healthcare^57^; urban land cover), *socioeconomic factors* (livestock density; relative social vulnerability), *ecosystem structure* (spatial vegetation heterogeneity as an indicator of landscape fragmentation; forest cover; cropland cover; biodiversity intactness index^58^), *land use change and intensity* (forest loss; cropland expansion; urban expansion; mining; protected area coverage; hunting pressure index) and *climate change* (mean change in annual temperature and precipitation between a 1950-1970 reference period and 2000-2020). These covariates were mainly derived from satellite remotely-sensed products, climate reanalysis and gridded demographic data, but some necessarily came from composite or modeled products, notably biodiversity intactness (average local abundance of all species relative to their abundance in minimally-disturbed habitat, predicted as a function of land use intensity^58^); social vulnerability (a composite indicator of relative multidimensional deprivation, based on inputs including infrastructure, human development index, nighttime lights and infant mortality rate^59^), and hunting pressure (estimated average hunting-linked abundance decline across all mammal species, predicted from several geographical covariates; tropical forest biomes only^60^). We used Bayesian logistic regression models to test the contribution of these covariates to outbreak risk, including continuous geospatial random intercepts to account for unexplained macro-scale patterns of data availability, for example between countries or subnational regions (Gauss-Markov random field, models implemented in INLA v23.3.26^61,62^; Methods, Extended Data Figure 2). Models were fitted to outbreak event records from 1985 onwards to align with the timescales of covariate data (with certain data-sparse exceptions that included data from post-1980; Methods). We apply this framework first to our entire dataset, and then on a disease-by-disease basis, and ask (1) whether a general anthropogenic fingerprint on the spatial intensity of outbreak events can be distinguished from both bias and noise; (2) whether any broad categories of environmental change are consistently implicated in outbreak events across pathogen types, transmission modes, and regions; and (3) whether there is evidence of widely-shared drivers across diseases and transmission modes that could point towards promising ecosystem-based intervention strategies.

## Detection biases shape outbreak hotspots at global and local scales

Globally, zoonotic, vector-borne, and environmentally transmitted disease outbreak events (n = 49,239 after data preprocessing, with 50,000 background points; Methods) were correlated with human-driven ecosystems (more forest cover, but more fragmented vegetation and lower biodiversity intactness) and less socially vulnerable communities (Figure 2a). However, these associations could be confounded by both broad- and local-scale biases in outbreak detection, investigation and reporting, as well as by the spatial extent of the specific sources that were compiled into our database. Extending the model to include a geospatial random effect and adjust for detection-related covariates showed that apparent hotspots are primarily created by these observation and reporting processes (Figure 2b-c). At its extremes, the magnitude of the geospatial effect exceeds all covariate effects, with the highest intensity in the United States and Brazil – two of the three countries whose national disease surveillance systems are substantially represented in our database – as well as in regional reporting hotspots across West and Central Africa, the Middle East, and South and Southeast Asia (Figure 2d). At a more local scale, outbreak events are much more commonly reported in cities and near clinics, with these two slope estimates much larger than any other covariate effects (Figure 2c). These relationships likely reflect the importance of health systems infrastructure in disease detection (and its direct influence on the location where outbreaks are reported), although our analysis cannot distinguish reporting bias from a true effect of urban environments on outbreak risk^63^.

**Figure 2:**
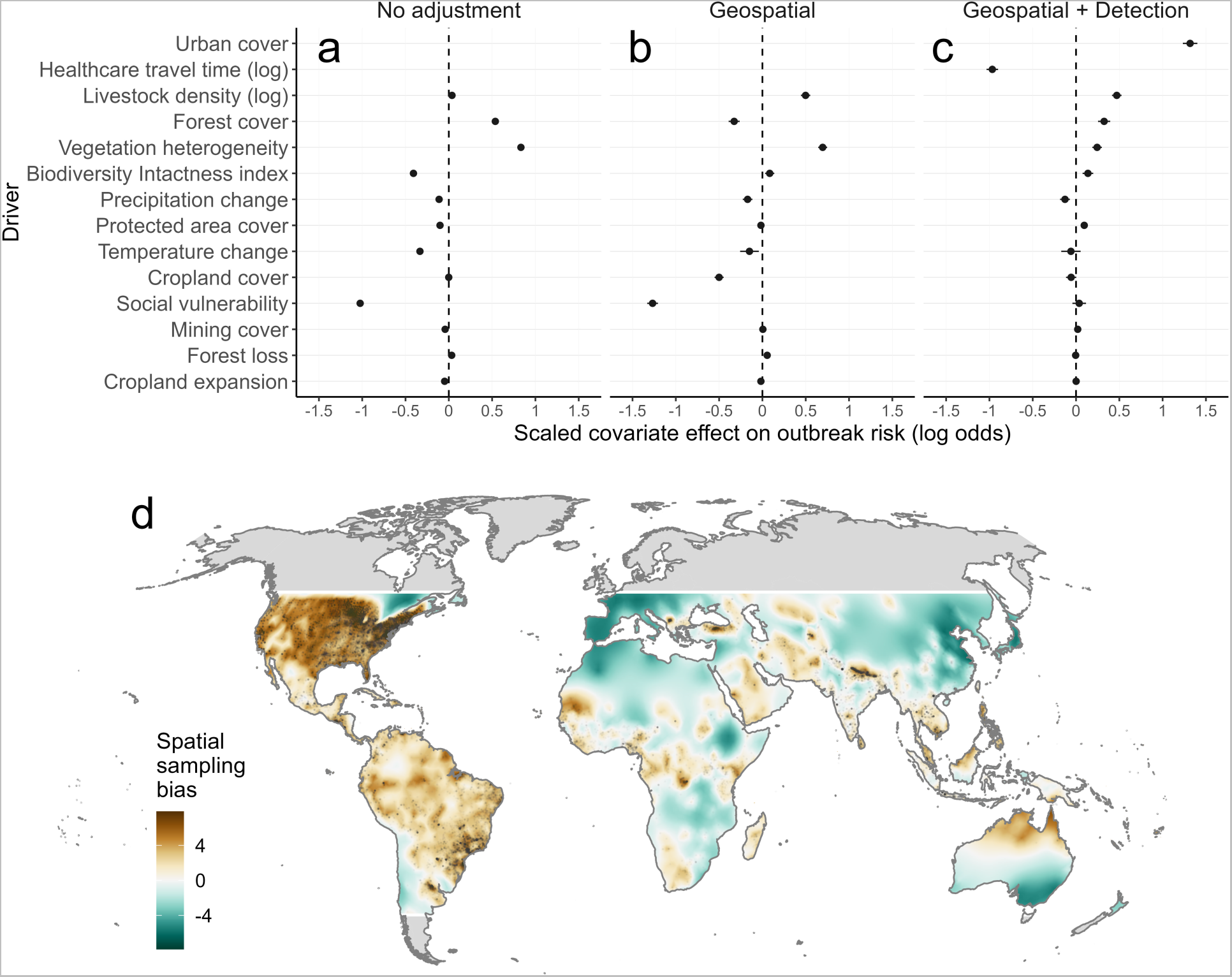
Geographical reporting and detection biases confound inference of global emerging infectious disease drivers. Points and error segments (A) show linear fixed effects of scaled covariates (posterior marginal mean and 95% credible interval) from Bayesian logistic regression models fitted to the full global dataset of contemporary outbreak events (i.e. pooling data across all diseases; n = 49,239 with 50,000 background points, between 1985-2022). Slope estimates denote the effect of each scaled covariate on spatial outbreak risk. Panels denote model specification: including either only socio-environmental fixed effects (*no adjustment*; A), adding a continuous geospatial random effect (*geospatial*; B), or adding a geospatial effect and local detection process covariates (*geospatial + detection;* C). The geospatial random effect (Gauss-Markov random field; D) reflects residual (unexplained) spatial variation in observed outbreak events, largely due to macro-scale sampling processes such as biases in awareness and reporting. Map color scale denotes contribution to the observed pattern of outbreak events (log-odds scale) with outbreak events overlaid as translucent points. The geospatial effect was only inferred within the latitudinal range of available data; areas outside these bounds are shaded in gray. Inferred effects were very similar for models fitted to subsets of diseases with different transmission characteristics (either zoonotic or vector-borne; Extended Data Figure 3).

Social and ecological risk factors were detectable, but with weaker or modified effects, after adjusting for detection and reporting processes (Figure 2a, 2c). Outbreak event risk was strongly associated with higher livestock density (especially when analysis was limited to zoonotic diseases, i.e. whose human infections arise principally from spillover from an animal reservoir; Extended Data Figure 3), forest cover and fragmented vegetation and, more weakly, with higher biodiversity intactness and protected area coverage. Outbreak events were also generally associated with areas experiencing long-term drying trends (Figure 2c). These results were very similar across models only including diseases classed non-exclusively as either zoonotic (n = 26 diseases) or vector-borne (i.e. transmitted by invertebrate vectors regardless of host type; n = 20) (Extended Data Figure 3). The findings of these global, disease-agnostic models (hereafter “global models”) thus broadly align with the consensus that emerging infectious diseases are associated with zones of frequent contact among people, livestock, and biodiverse ecosystems^1,4–6^. However, spatial reporting biases at multiple scales, both regional and highly localized, have by far the strongest influence on the inferred global geography and drivers of outbreak events. These biases, and the socio-ecological diversity of disease systems represented in our dataset, emphasize the need to transition towards more granular (and bias-adjusted) disease system-specific inference.

## Anthropogenic drivers of outbreak risk are detectable and differ across disease systems

Next, we developed disease-specific geospatial models for 31 diseases (excluding Hendra virus disease, due to data sparsity [n = 11 outbreak records]). The high number of pairwise disease-driver combinations (n = 496) and spatial reporting biases created a substantial risk of detecting spurious relationships. Therefore, to ensure we only tested specific and plausible hypotheses, we conducted a participatory hypothesis-generating exercise in which 25 study authors independently ranked candidate drivers for each disease (Supp. Table 2; Methods). The results were used to identify specific drivers to test per disease, based on either broad or strict thresholds for consensus (Methods, Extended Data Figure 4). Overall, the top-ranked hypothesized drivers were healthcare access and socioeconomic vulnerability, followed by landscape fragmentation, deforestation, urbanization, and climate change-related variables (Extended Data Figure 4). We fitted hypothesis-driven multivariable models for each disease following the general methods described above, with geospatial effects and both detection process covariates included in all models as *a priori* expected confounders (except when detection covariates were highly collinear; Methods, Extended Data Figure 3). We also fitted univariable models, i.e.each driver individually plus a geospatial random effect, to compare to inferred effects without adjustment for local detection covariates (Extended Data Figure 5).

Across diseases, we again found widespread evidence of systematic reporting biases: the most prevalent significant predictors were increasing urban land cover (20 out of 30 diseases tested) and proximity to the nearest health facility (15 out of 23 tested); these predictors also had the two largest average scaled effect sizes (Figure 3, Extended Data Figure 6, Supp. Figure 1). For approximately half of the diseases we examined, we again found that outbreak risk was higher in fragmented and forested landscapes, with common and almost always positive effects of vegetation heterogeneity (15 out of 30 tested) and forest cover (13 out of 27 tested) (Figure 3). For a quarter of the diseases we examined (8 out of 31 tested), outbreak event risk was higher in localities experiencing long-term changes in annual precipitation: in particular, climate drying was strongly associated with several vector- and water- borne diseases with known or suspected links to anomalous drought-wetness dynamics (e.g. Rift Valley fever^64^, dengue fever^65^, melioidosis^66^ and Japanese encephalitis^67^; Extended Data Figure 5).

**Figure 3:**
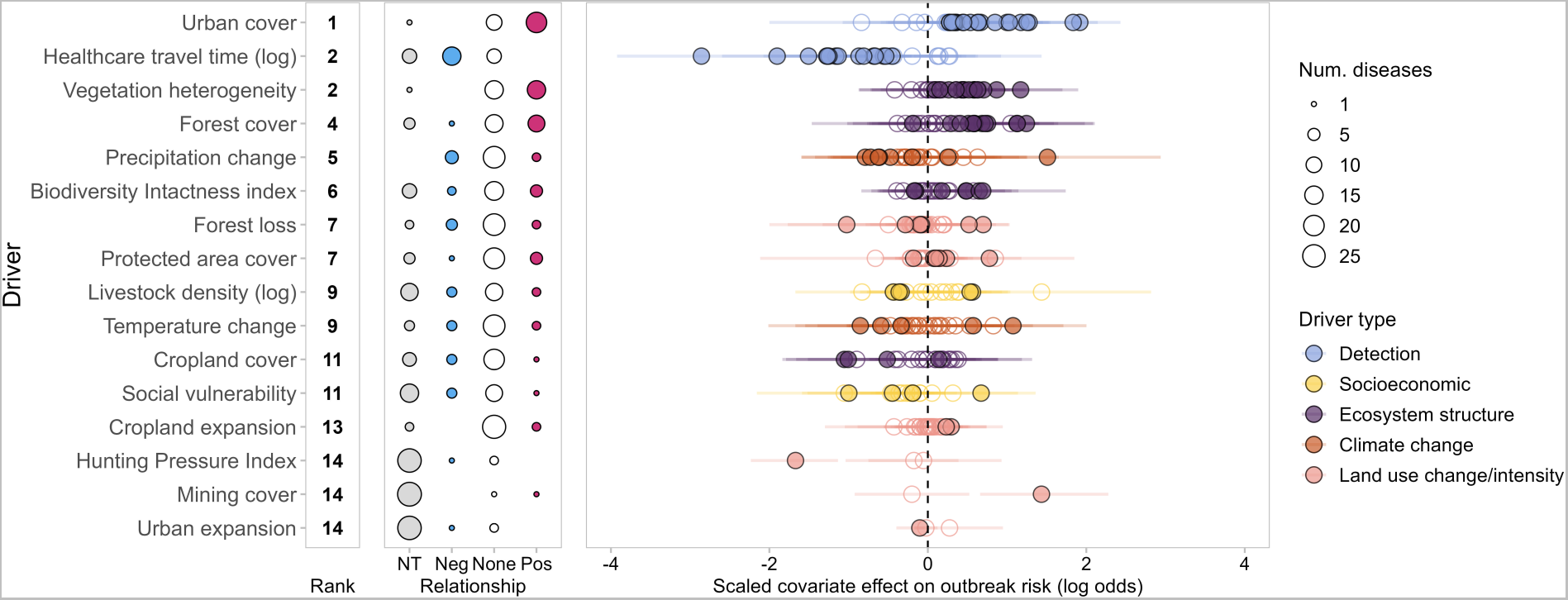
Drivers of outbreak risk across 31 emerging infectious diseases. Results are summarized from separate hypothesis-based geospatial logistic regression models for all 31 diseases in the dataset (Extended Data Figure 5). Drivers are shown ranked by the number of diseases for which there was strong evidence of a relationship (95% credible interval not overlapping zero) (left column). The prevalence and directionality of driver effects is summarized for each driver (middle column), with point size showing the number of diseases for which relationships were either not tested (*NT*; gray), negative (*Neg*; blue), positive (*Pos*; red), or no strong evidence (*None;* i.e. 95% credible interval overlapping zero). Points and error segments (right column) show driver fixed effect parameters on the log-odds scale (posterior marginal mean and 95% credible interval) for all tested diseases, with filled points denoting strong evidence of a relationship, and point color denoting the broad class of socio-environmental driver. Results are based on hypotheses generated using a broad “majority rule” criterion; results for stricter models testing only top-ranked drivers per-disease are qualitatively very similar (Extended Data Figure 6).

Relationships with land use, biodiversity, temperature, and socioeconomic factors were less commonly detected across diseases, more heterogeneous in effect size and direction (Figure 3; Extended Data Figure 5), and often failed to align with their hypothesized importance (Extended Data Figure 4). Although deforestation is often considered one of the most common drivers of disease emergence, we detected impacts of recent cumulative forest loss (2000-2020) in one quarter of systems for which this driver was tested (7 out of 29 tested), with a mix of directional effects (three positive, four negative) and small effect sizes (Figure 3). There were similarly varied relationships with biodiversity intactness, protected area coverage, and livestock density (respectively 7 out of 24; 6 out of 27; and 5 out of 18 tested; Figure 3), the latter contrasting notably with the large positive effect of livestock density in the global model. Warming was a hypothesized driver for most diseases, but we only detected effects of long-term temperature change for a few diseases (5 out of 28 tested) again with little consistency in direction (three positive, two negative; Figure 3). Finally, although social vulnerability was one of the highest-ranked hypothesized drivers, disentangling any signal from detection biases proved impossible at this broad scale (and with a relatively coarse global vulnerability indicator): outbreak events were strongly biased towards more affluent settings in univariable models (25 out of 30 diseases tested), but these effects almost always became negligible after adjusting for detection covariates (Extended Data Figure 5). All of these findings were very similar when testing hypotheses generated using a stricter criterion for consensus (Extended Data Figures 5-6).

It is unclear to what degree the limited detectability of certain drivers reflects a true absence of causal relationships, or is primarily a byproduct of data sparsity, spatial misalignment between where infections occur and where they are detected, temporal misalignment between infections and environmental driver data, and/or measurement error in the environmental driver covariates (Methods). Because of these limitations, our framework may not always detect weaker, more confounded or time-sensitive effects (e.g. transient changes in risk during the land conversion process^30^), especially for data-limited diseases. To some degree, these limitations may be inherent to cross-disease geospatial analyses at continental or global scales; we therefore suggest our approach should be thought of as complementary to system-specific work, including both longitudinal eco-epidemiological studies^7^ and ethnographic research^40^. Nonetheless, our confidence in the overall findings was strengthened by a sensitivity analysis of arbovirus surveillance data from the United States, which showed that our outbreak event case-control framework can detect similar spatial drivers as full models of county-level case incidence (Extended Data Figure 7). Our models also detected numerous well-known or strongly-suspected drivers of specific diseases, further validating the approach: these included a negative effect of biodiversity intactness on Lyme disease (consistent with foundational disease ecology research into the dilution effect^8,68^); pig density and both temperature and precipitation change trends as drivers of Japanese encephalitis^69^; an increased risk of avian influenza A/H5N1 outbreaks in areas with higher poultry densities^70^; positive impacts of forest loss on mpox and zoonotic malaria^71^; and evidence of fragmented forest cover driving human outbreaks of arboviruses that emerge at human-forest ecotones^72^ (i.e., Mayaro fever, Oropouche fever, and yellow fever) (Extended Data Figure 5). Finally, for Argentine hemorrhagic fever, after adjusting for detection processes we found evidence that outbreak events are more frequent in relatively more socially-vulnerable areas, which aligns with the disease’s rodent-borne transmission ecology^73^. Surveillance data on this disease were the most consistently and precisely geolocated in our entire database (Extended Data Table 1), demonstrating the value of precise, standardized spatial disease surveillance reports for reducing the confounding impacts of detection bias.

## Shared drivers and syndemic risks differ by pathogen transmission mode

Prevailing narratives about disease emergence tend to focus on the impacts and relative importance of individual drivers, but outbreak risks often arise through synergistic interactions between diverse socio-environmental processes^7,74^. For pathogens with shared ecological characteristics, convergence of socio-ecological drivers - for example, similar vector community responses to land use pressures^29^ - might produce clustering of multiple infections within the same population, potentially leading to worse outcomes (“syndemics” or syndemic interactions)^74–76^. Differences in the landscape-level structure of anthropogenic impacts could even help to explain global syndromes of disease emergence: for example, in East Asia and the Pacific, most drivers we analyzed are tightly correlated across space, while the opposite is true in sub-Saharan Africa (Extended Data Figure 8).

To explore how these kinds of interactions could affect outbreak risks, we visualized patterns of driver occurrence and co-occurrence as unipartite networks (Figure 4, Extended Data Figure 6), across all 31 disease-specific models and separately for either directly transmitted (n = 10) or vector-borne zoonoses (n = 16). We found that certain drivers co-occur frequently overall – principally urban cover and healthcare access, and to a lesser extent fragmented vegetation and forest cover (Figure 4a) – and that this pattern does not simply reflect landscape structure; for example, forest cover and vegetation heterogeneity are uncorrelated globally, while cities, travel time to healthcare, and vegetation heterogeneity are at most moderately correlated (Extended Data Figure 8).

**Figure 4:**
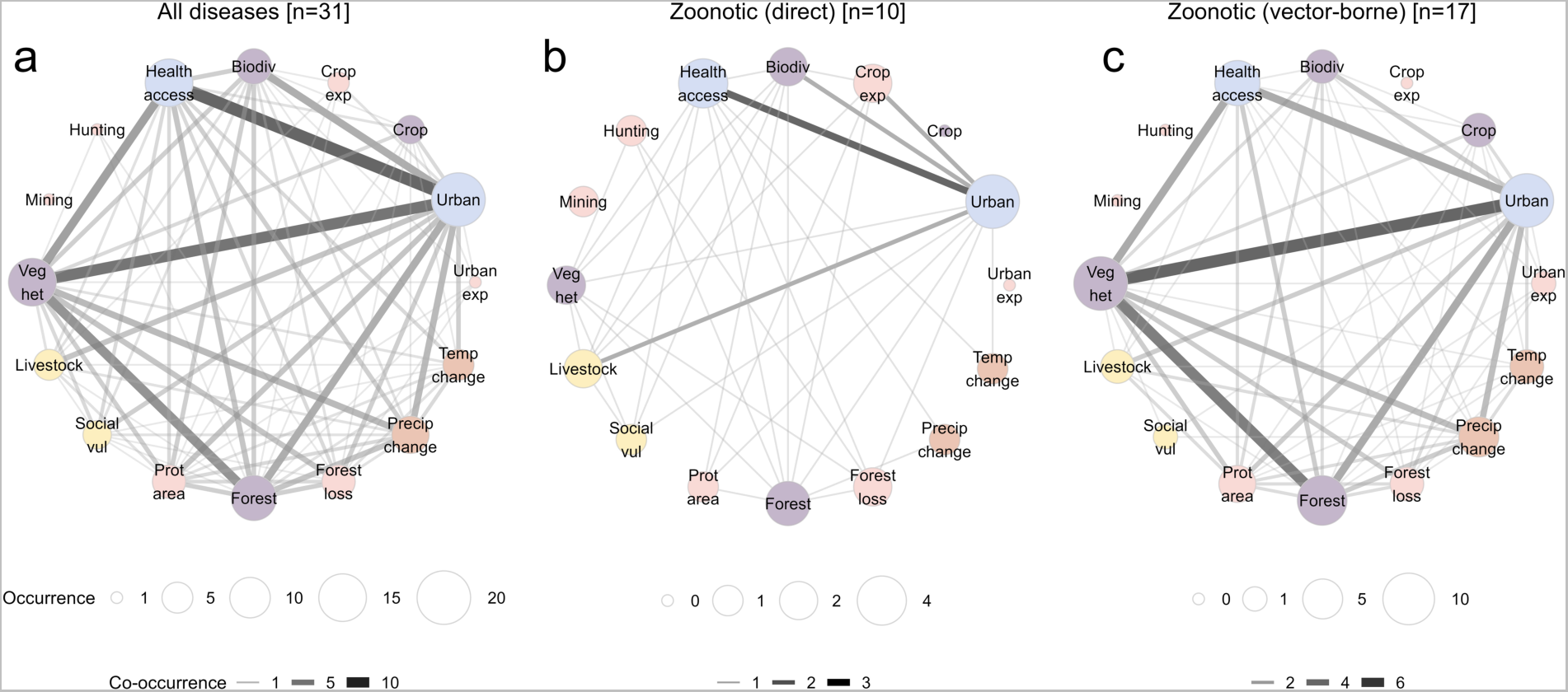
Co-occurrence of outbreak drivers across emerging infectious disease groups. The patterns of co-occurring drivers across individually-modeled diseases are represented as unipartite networks, for all diseases (n = 31; A) and for subsets of zoonotic diseases whose predominant mode of transmission to humans is either direct (B; n = 10) or vector-borne (C; n = 17). Nodes represent drivers, with node size proportional to the number of diseases with strong evidence of a non-zero effect (“driver occurrence”), and edge weight is proportional to the number of diseases for which driver pairs co-occur (“driver co-occurrence”; i.e., non-zero inferred effects of both drivers in multivariable models). Node colors denote driver type, as in Figure 3. Larger nodes reflect more prevalent drivers and darker edges reflect a higher prevalence of driver co-occurrence within each group of diseases. Results are derived from multivariable models testing hypotheses under the “majority rule” criterion; the pattern of clustered drivers is the same under the stricter “top ranked” criterion (Extended Data Figure 6). The pattern of co-occurring drivers does not simply reflect observed correlation between variables, as many of these variables are weakly or uncorrelated globally (Extended Data Figure 8).

Notably, patterns of driver co-occurrence differ substantially by transmission route. In particular, zoonotic diseases that transmit from animals to humans through an arthropod vector (e.g., Chagas disease, Lyme disease, or yellow fever) have a proportionally higher rate of co-occurring ecosystem drivers (Figure 4c). This reinforces existing evidence about vector-borne disease risks in degraded and urbanizing landscapes^28,77,78^, particularly where compound drivers and shared vectors (e.g., *Aedes* mosquitoes) could interact to create syndemic risks^79,80^; and suggests that ecosystem-based strategies (such as protecting intact forests, or regulating the financial actors most responsible for unsustainable, extractive land use^81^) may be effective in mitigating the burden of these diseases.

In contrast, we found little evidence of widely shared spatial drivers among directly transmitted zoonoses (e.g., Ebola virus disease, MERS, or mpox) (Figure 4b). This may be due in part to the relative paucity of outbreak data for several of these pathogens, which constrains inferential power (Figure 1b-c), but it also likely reflects their diversity of ecologies, life cycles, and human exposure pathways (e.g., hunting, contact with livestock and food products, household contact with wildlife^82^, or occupational contact with wildlife, such as through agriculture^83^). These findings do not support the idea that one-size-fits-all ecological interventions (e.g. tighter global regulations on deforestation and agricultural expansion) would be broadly protective against epidemic and pandemic threats, such as directly-transmitted respiratory and hemorrhagic fever viruses. Ecosystem-based risk prevention and surveillance programmes remain the most effective and scientifically-supported option to reduce spillover risk and improve outbreak detection at the human-animal interface^16^, but our findings suggest that proposed interventions should be tailored to the ecology of specific priority pathogens in specific landscapes. In systems where this evidence is currently limited, long-term ecological research can establish these principles in striking detail^7^.

## Healthcare access supports both surveillance and response

Even with our extensive dataset of 32 diseases, representing a wide variety of different pathogens, biomes and socioeconomic contexts, our study remains limited by sample size and data quality. Detection and reporting biases are a pervasive, worldwide phenomenon, spanning multiple spatial scales and low- to high-income settings (Figures 2 and 3). Strikingly, many of the highest-concern diseases – such as bat-borne epidemic viruses – have the lowest availability of data (Figure 1). These gaps probably reflect under-detection rather than a true scarcity of spillover events: previous studies have estimated that up to half of all Ebola outbreaks might never have been identified^84^, a pattern that serological evidence indicates also applies to many high-concern zoonotic pathogens (e.g. SARS-related bat coronaviruses^85^ and Lassa fever^86^).

These findings highlight an underappreciated and disease-agnostic lever for intervention: improving access to healthcare in underserved rural and remote communities. In much of the world, it can take over a day to reach a clinic, especially without motorized transportation^57^, and remote clinics often lack capacity for molecular diagnostics, especially for rare infections; these gaps in health systems are likely to be persistent at high-risk interfaces between rural communities and intact ecosystems. Outbreaks that start further from clinics are less likely to be detected, promptly diagnosed and treated, and – without a timely response – may be more likely to grow into epidemics^87^. For most of the diseases we examined, outbreak event reports are clustered in close proximity to clinics: outbreak odds declined by a median of 32% (range 1.2%-96.7%) for each additional hour’s motorized travel time from the nearest healthcare facility (Figure 5a). Average travel times to healthcare across each disease’s entire geographic range are generally much higher than at documented outbreak locations, with a substantial proportion of population-weighted background locations falling over 2 hours away (median 23%, range 2%-38%; Figure 5b-c). These travel time estimates may also be relatively conservative, given socioeconomic disparities in access to motorized transport, the tendency for models to underestimate actual travel times (e.g. due to road quality or traffic)^88^, and the many additional non-geographic barriers to accessing healthcare. Investing in new infrastructure and lowering social and economic barriers to access would ensure timely disease diagnosis, treatment, and prevention for underserved communities — and would substantially increase the odds of outbreak detection and reporting. Improving global surveillance of infectious diseases at human-nature interfaces would also help address the data gaps highlighted in our study, and could therefore help strengthen the scientific evidence base around ecological strategies for risk reduction.

**Figure 5:**
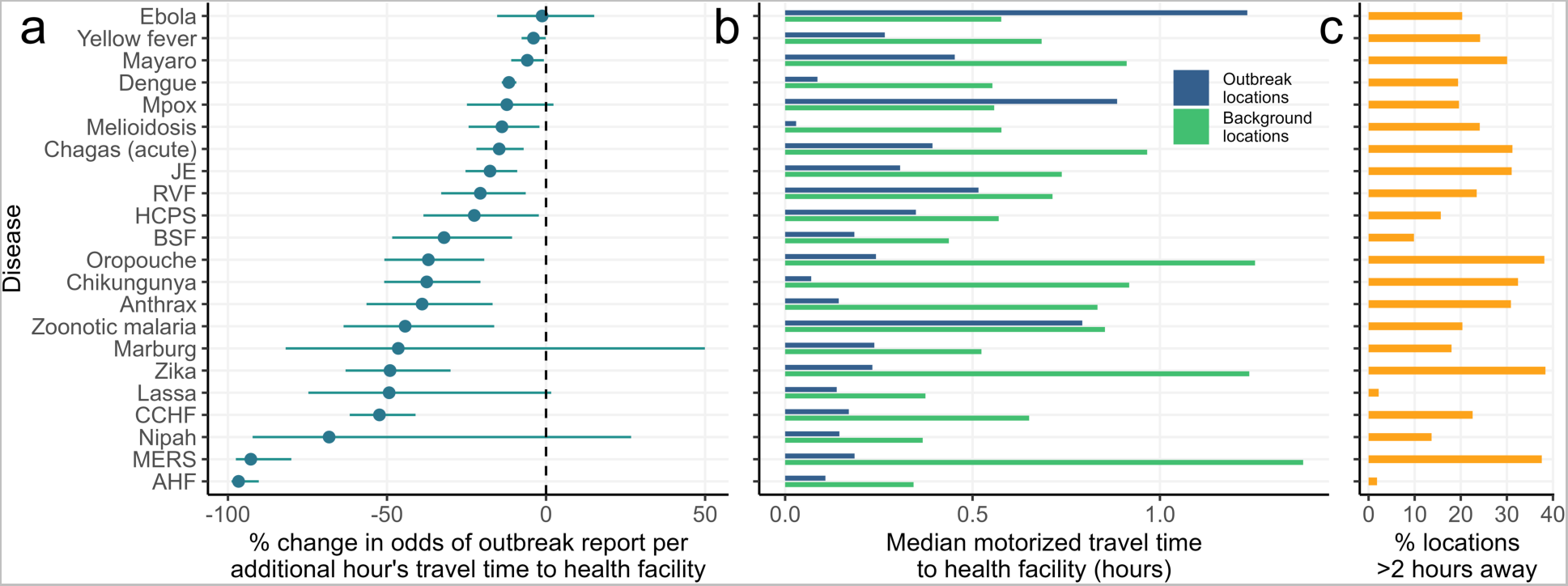
Outbreak detection declines with increasing distance from healthcare facilities. Using the inferred slope parameters from each multivariable disease model, we estimated the marginal percentage change in odds of outbreak reporting for each additional hour of motorized travel time from the nearest health facility (A; points and error segments show posterior median and 95% credible interval). Barplot (B) shows, for each disease, the median motorized travel time to the nearest health facility across outbreak locations (blue) compared to population-weighted background locations (i.e. a representative background sample across the at-risk area; green). Barplot (C) shows the percentage of population-weighted background locations falling more than 2 hours from the nearest health center. This covariate effect was tested for 23 diseases (22 shown; St. Louis encephalitis was not visualized due to extremely wide uncertainty) but not for the remaining 8 diseases (mostly in the US) due to high collinearity with urban cover (Methods).

## Conclusion

The recent rise of emerging infectious diseases is often described – both by scientists and science communicators – as a byproduct of global anthropogenic environmental change^20,91^. This trend may share common causes with both the climate and biodiversity crisis, but is also a “wicked problem” in its own right: our analyses suggest that emerging infectious disease risks are ubiquitous, and widely associated with mosaic landscapes where people and cities live alongside forests and fragmented ecosystems. For many vector-borne diseases, we find evidence of a strong and consistent anthropogenic fingerprint, supporting the idea that certain land use and climate policies could achieve net reductions in disease burden. However, we were unable to detect a similarly consistent anthropogenic fingerprint on directly-transmitted zoonotic infections, diverging from popular emergence narratives that have been based largely on evidence from in-depth case studies^2,91,92^. In any given region, investments in ecological and community-led research will be needed to identify and evaluate locally-tailored ecosystem interventions that reduce spillover risk and the endemic burden of regional priority diseases. Meanwhile, as new diseases continue to emerge – and human activities continue to transform the planet, even in the best-case scenarios for sustainable development – we suggest that the global community should redouble their investments in health system strengthening. Achieving universal health coverage, strengthening outbreak response capacity, and investing in novel vaccines and therapeutics, can help to ensure that - even in a world with 5% more spillover events each year^93^ – outbreaks never have the opportunity to become epidemics.

## Materials and Methods

### Overview

The aim of this study was to empirically test for a general detectable fingerprint of anthropogenic drivers on the geographical distribution of human outbreaks of 32 emerging infectious diseases, based on existing geolocated outbreak and case data sources, as well as gridded datasets representing key socio-environmental disease drivers. To account for differing ecological characteristics across diseases and avoid testing for spurious or irrelevant associations, we generated a set of hypothesized key drivers for each individual disease through a structured form-based exercise completed by most coauthors, whose expertise spans a wide range of disciplines and scales of enquiry (from microbiology to global public health). Across all diseases overall, and individually per disease, we applied a standardized statistical inference framework, which involved harmonization of point and polygon data and inference of the drivers of outbreak risk using geospatial logistic regression models. We describe these methodological stages in detail in the following sections.

### Collection and harmonization of geolocated human disease data

We collated and harmonized geolocated point and polygon data on human case occurrence and/or incidence of 32 environmentally-linked emerging infectious diseases, from numerous published datasets in the scientific literature and from open national disease surveillance data portals (Figure 1, Extended Data Table 1, Supp. Table 1). We used the following broad criteria to select diseases for inclusion: (1) Human infection risk should be closely coupled and thus in principle attributable to local environmental or ecological conditions (i.e. zoonotic, vector-borne, or environmentally-transmitted), and if extended human-to-human transmission chains independent of these conditions are possible, datasets must specify the locations of probable index cases. (2) Diseases should not be sufficiently well-surveyed that prevalence surveys, rather than case incidence or occurrence, could form the basis for inference. (3) Diseases should not have been subject to long-term eradication programmes that could confound inference of environmental drivers. These criteria meant that our analyses included many emerging, rare and high-concern zoonotic and vector-borne pathogens (including many mosquito-borne arboviruses, rodent- and bat-borne viruses and *Plasmodium knowlesi* zoonotic malaria), but not *P. falciparum* or *P. vivax* malarias or neglected tropical helminthiases.

The full list of diseases, data sources and their spatial and temporal coverage is provided in Extended Data Table 1. When compiling data for each disease, our priority was to select datasets that covered as much of the known geographic extent of transmission as possible, while remaining internally consistent (i.e. collated in a standardized and comparable way to facilitate analysis). We focused on compiling existing published datasets from scientific literature and openly accessible disease surveillance portals, rather than collecting additional data (e.g. via scraping scientific literature or ProMED), to ensure that our analyses are representative of data that are currently in the public domain and relatively analysis-ready. Notably, sufficient or suitable data were not available for certain high-priority diseases, most notably SARS-related coronaviruses, because too few confirmed spillover events have been documented to provide a geographic picture of risk^94^. Datasets were obtained either via downloading from scientific paper supplementary data or open repositories, sharing between study coauthors, or through email requests to specific paper lead authors. To credit the substantial work involved in compiling the source datasets and ensure our author team included disease-specific expertise, lead authors who collated and shared datasets were invited to be study coauthors and participate in hypothesis generation (see below) and manuscript writing and editing (see Author Contributions section for a breakdown of roles).

Human case datasets are generally available in one of two formats. *(1) Geolocated spillover or outbreak occurrences.* Here, records represent 1 or more cases occurring at a named place and time, with geographical precision ranging from a specific point location or point with buffer radius (more precise), to a named administrative unit (less precise). This category of data includes most of the datasets collated for the purpose of risk mapping, for example by research groups affiliated with the US-based Institute for Health Metrics and Evaluation^53,95^.*(2) Case counts from named areal units.* Here, records contain the number of cases reported from a particular areal unit (usually administrative level 1 or 2) during a particular time window (usually month or year). This category mainly includes datasets collected and reported through national notifiable disease surveillance systems, which are often available via online portals, reports, or scientific papers. Point locations can provide greater geographic precision on environmental conditions nearby to a reported disease case, whereas administrative units require averaging conditions across often much-larger polygons. Consequently, different sources provide different levels of information about both transmission intensity (binary outbreak occurrence versus number of cases) and environmental context (specific event location versus broad aggregated unit).

To ensure that the results of our models were comparable across diseases and datasets (Extended Data Table 1), we therefore needed to develop a common harmonization framework to accommodate these diverse data sources while preserving spatial uncertainty in location of infection. A diagram of this pipeline is shown in Extended Data Figure 1 and described as follows. The response variable, an “outbreak event”, was defined as at least 1 case in a named locality in a given year, to ensure comparability in analyses between geolocated outbreak datasets (which contain no or partial information about the number of cases) and surveillance data (which typically provide an estimate of incidence). For any given disease, all outbreak locations (whether natively point or polygon) were converted to polygon objects using the ‘sf’ package in R^96^, by drawing a circular buffer around point locations with a radius of either 5km or another custom value (if specified within the source dataset). All polygons covering too large a spatial area were excluded as too imprecise to link to local environmental conditions; this was by default >5000 km^2^ (equivalent in area to a circular buffer with a radius of 40km) but was relaxed to higher values (mostly under 10,000km^2^, but maximum 20,000km^2^) for certain data-sparse diseases and coarser areal case surveillance datasets (Brazilian spotted fever, chikungunya, Eastern equine encephalitis, influenza (H5N1), Jamestown Canyon encephalitis, Marburg virus disease, Mayaro fever, Oropouche fever, plague, Rift Valley fever, St. Louis encephalitis, West Nile fever, and yellow fever), as a compromise to retain as complete a geographical picture of outbreak event distributions as possible.

For each disease, this process produced a dataframe where each row with a unique identifier represents an outbreak event (i.e. 1 or more cases in a given locality in a given year) with metadata where available (number of cases, case definition, diagnostic method, etc), along with an associated shapefile linking each record to a geographical polygon. For most diseases the shapefile contained a mixture of smaller circular buffers around point locations (with radius between 5 and 20km) and larger, irregularly shaped administrative unit polygons. Across all diseases, the full database contained 58,319 unique georeferenced outbreak events, for 32 diseases, in 169 countries worldwide (Figure 1). The majority (88.7%) of records were from after 2000, whereas far fewer records (2.4%) were from before 1980. The constraints of available data mean that these datasets are necessarily presence-only (i.e. contain only information on positive case detections without true negatives as controls), so later modeling analysis required the selection of background points as pseudo-controls (Extended Data Figures 1-2); we describe this process below.

### Disease-specific hypotheses for the drivers of human infection risk

A large body of literature has proposed that several broad anthropogenic change processes may be general or common drivers of risk across a large number of zoonotic, vector-borne and environmentally-mediated diseases (e.g. agriculture and urban expansion, deforestation, wildlife hunting, biodiversity loss). Yet given the wide diversity of reservoir hosts and transmission ecologies across pathogens, spatial drivers of risk may often be pathogen- or context-specific. To ensure our analyses accounted for expected ecological differences between systems, we developed a structured, form-based exercise to identify key hypothesized drivers for each individual disease, which was then completed by study coauthors. To balance between comprehensiveness and exhaustion, we created a fill-in matrix spreadsheet of 18 drivers and 34 disease systems (see Supp. Table 2). Each cell could be filled in by the respondents indicating their choice of a driver having a negative, positive, none, or ‘don’t know’ impact, and respondents were additionally asked to provide a 1-3 ranking for their expected top three drivers for each disease (in either direction). Given that coauthors have a range of expertise, which may include either multiple disease systems, or a focus on one or a few, respondents could choose to leave one or more full disease systems blank (NA).The full list of drivers included ecological/environmental processes (biodiversity loss, forest cover, forest loss, invasive species, long-term temperature change, long-term precipitation change), processes driving human-wildlife contact (cropland cover and expansion, landscape fragmentation, mining, protected area coverage, urban cover and expansion, wildlife hunting, wildlife trade and markets) and social processes influencing exposure and detection (socioeconomic vulnerability, proximity to hospitals/clinics, livestock density). This hypothesis-generation exercise was intended mainly to robustly identify a group of testable drivers for each disease that reflect system-specific knowledge, but this process also allowed us to compare between coauthor opinion and what can be inferred from available data.

This exercise was completed by most coauthors (25 of 31; Extended Data Figure 4), whose expertise spans multiple disciplines, disease systems and scales of biological organization, including microbiology and virology, genomics, disease ecology and evolution, epidemiology, veterinary medicine, social-ecological systems, public health, and machine learning and statistical inference. Despite this wide disciplinary expertise this group still consists largely of academic researchers based in Global North institutions, and as such our hypotheses are unlikely to fully reflect locally-situated understandings of most of these diseases. While this exercise was a concise approach to hypothesis generation, users reported spending multiple hours (>2) to fully complete the matrix; this is feasible and reasonable for invested author teams such as this, but we do not recommend this as a template for a rapid, large-scale survey exercise.

We then post-processed the completed exercise data to generate hypothesized drivers for each disease using two definitions of group consensus. For each disease we first adopted a broad definition, including drivers for which more respondents stated an effect (either positive or negative) than stated no effect (“majority rule”). As a sensitivity check we also adopted a narrower definition, including only drivers that were included in the top 3 ranked drivers by at least 1 respondent (“top-ranked”). This process generated consensus lists of hypothesized drivers to test for each disease (Extended Data Figure 4) reducing the risk that models would identify spurious, ecologically-implausible drivers (e.g. wildlife hunting for West Nile). Such an issue could otherwise feasibly arise due to the small size and spatially-biased nature of many disease datasets (see “Limitations of data and methodology” below).

### Collation of geospatial data on socio-environmental drivers of disease

In parallel, we collated global geospatial (raster) layers describing socio-environmental and climatic features as proxies for the key geographic drivers of risk listed above, based on remote sensing, climate reanalysis, social indicators and census-based data sources. A full table of socio-environmental covariates, their sources and processing is provided in Extended Data Table 2 and Supp. Table 3. The small size of many disease datasets unfortunately meant there was insufficient data to analyze the relationship between cases and covariates in both space and time, which therefore limited our study to spatial rather than spatiotemporal driver analysis (see “Limitations” below). Therefore, for variables describing gross characteristics of the environment (e.g. land cover type proportion variables) we selected a single raster year or time period close to the central tendency of reported disease data (i.e. between 2005 and 2015), while aiming for the best spatial and thematic resolution possible under that constraint. For variables describing anthropogenic change, we generated rasters that described the grid cell-level change in a particular variable across most of the disease data period (e.g. tree cover loss between 2000 and 2020, change in mean annual temperature between 1950-1970 and 2000-2020). Raster covariates were used at their original spatial resolution with a few exceptions (e.g. social vulnerability was aggregated; see Extended Data Table 2); since this was not a mapping study no rescaling was required. Notably, we were unable to identify suitable proxy covariates for several widely-hypothesized drivers that have not been quantified in space and time, highlighting an important lack of systematic data collection around key putative drivers of disease emergence; these include invasive species density, wildlife trade and/or live markets, and wildlife hunting outside tropical forests.

A brief description of the full list of the socio-environmental raster datasets is as follows: *temperature change* (change in grid-cell level mean annual air temperature between reference period of 1950-1970 and focal period of 2000-2020, derived from ERA5-Land reanalysis^97^); *precipitation change* (change in grid-cell level mean annual precipitation between 1950-1970 and 2000-2020, from ERA5-Land); *forest cover* (grid cell-level fractional tree cover from Copernicus land cover 2015); *forest loss* (grid cell-level tree cover loss 2000-2020 from Global Forest Change); *biodiversity intactness* (local Biodiversity Intactness Index, modeled for 2005 based on human disturbance layers^58^); *cropland cover* (grid cell-level fractional crop cover from Copernicus land cover 2015); *cropland expansion* (grid cell-level cropland growth 2000-2019^98^); *vegetation heterogeneity* (grid cell-level EVI dissimilarity index 2005, a metric of landscape fragmentation sensitive to anthropogenic landscapes^99^); *hunting pressure index* (a modeled defaunation index measuring average hunting-related species declines in tropical forest biomes^60^); *protected area cover* (whether grid cell is under area-based conservation, based on the World Database of Protected Areas 2022); *mining cover* (whether grid cell is under mining land use, based on ref.^100^); *social vulnerability* (the Global Gridded Relative Deprivation Index for a nominal present-day period^59^); *travel time to healthcare* (road-based travel time to nearest hospital or clinic for nominal year 2015^57^); *urban cover* (grid-cell level fractional urban cover from Copernicus land cover 2015); *urban expansion* (grid-cell level expansion of built-up areas 2000-2019 derived from ESA-CCI land cover); and *livestock density* (grid-cell-level density of livestock types from Gridded Livestock of the World v3).

### Statistical modeling

To infer the drivers of the geographic distribution of human cases while accounting for spatial and detection biases, we applied a standardized geospatial modeling approach for each disease. We describe this procedure in the following paragraphs.

For each model, we first defined the geographical boundaries of the modeling area (“study region”). For datasets compiled from the scientific literature, this was defined as a smoothed convex hull polygon around the full extent of geographical case occurrences, with a buffer of 180km (Extended Data Figures 1-2). For national-level case surveillance data the study area was constrained to the borders of the relevant country or subnational region. We then generated a final case-control dataset for modeling. We excluded records from before 1985 for most diseases, to better align the disease data with the timescale of available covariates; exceptions were certain data-sparse diseases where data from post-1980 were kept to retain as much information as possible (anthrax, Ebola virus disease, Marburg virus disease, Mayaro fever, and Oropouche fever). Because the case data were presence-only, meaning there were no true negative controls, we then generated background (pseudo-control) points throughout the study region. We selected between 2 and 8 times as many background points as presence points; this varied depending on the number of positive observations and their geographical dispersion, with higher multiples selected for more widely-distributed but data-sparse diseases to capture the full background area. (Guidelines have been developed for the selection of background points for species distribution modeling, which often lean towards balanced training sets of presence and pseudoabsence points^101^, but we note that this is a distinct statistical approach; our objective here is to detect predictor effects, rather than correctly model the area of occupancy, and as such our priority is statistical power and coverage of the area being examined.) All else being equal, the null expectation is that the distribution of human disease cases would follow the distribution of population; as such, entirely spatially random selection of background points would over-represent sparsely populated rural areas and under-represent highly populated urban areas. We therefore weighted background points distribution by human population, i.e. randomly generated point locations with the probability of a location being selected proportional to log+1-transformed population. This was based on a global raster of 2010 human population per pixel (WorldPop’s top-down unconstrained mosaics^102^), at 1km resolution for most diseases, but 10km resolution for certain diseases spanning a multi-continent geographic range to limit computation time (e.g. dengue, chikungunya). This approach produced a pseudo case-control design, i.e. comparing the socio-environmental conditions experienced by human populations at the locations where outbreaks have occurred (cases), to a representative background sample of the conditions experienced by populations across the study region (“controls”). Circular buffers were created around each background point with an area equal to the median area of the outbreak location polygons, to ensure covariates were averaged across a comparable geographical area for both presence and background points (Extended Data Figure 1).

For each model, this process produced a final dataframe of presences and pseudo-absences with associated polygons (again using ‘sf’), from which we extracted the mean value for each raster covariate using the ‘exactextractr’ package. We excluded from the analyses any variables that were missing data for >10% of observations or contained zeroes for >95% of observations. We examined collinearity among covariates via visual inspection, correlation matrix plots and variance inflation factors, and identified and excluded highly collinear covariates from multivariable models; this step was conducted manually rather than programmatically, to prioritize the inclusion of covariates with a strong hypothesized relationship to each disease in question. The final sets of covariates included in each disease-specific multivariable model are visualized in Extended Figure 5.

To infer relationships between covariates and disease outbreak probability 𝑝 at location 𝑖 (log odds of occurrence), we fitted geospatial logistic regression models in a Bayesian inference framework (integrated nested Laplace approximation, implemented in the package ‘INLA’ v23.3.26^61,62^), with the following general formula:

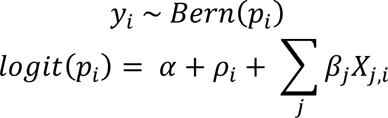

Here, 𝛼 is the intercept, 𝜌_!_ is a continuous spatially-structured random effect, and 𝛽 is a vector of linear fixed effects parameter estimates for the matrix of 𝑗 covariates 𝑋_*j*_. The geospatial effect was specified as a Gauss-Markov random field fitted using a stochastic partial differential equations approach (SPDE), with penalized complexity priors on the range and sigma parameters, and an intermediate mesh density chosen to reasonably balance between spatial precision and computation time. We set Gaussian priors for intercept and linear fixed effects (mean = 0, precision = 1). Due to wide variation in geographic range size and patchiness of data across the different diseases, the hyperparameters of the SPDE model’s Matern covariance function (range and variance) were manually tuned for each disease to ensure a smooth fit of the spatial field, assessed via visual inspection. After model fitting, we visually inspected posterior parameter and hyperparameter distributions, visualized the fitted SPDE to check for any visible issues with inference of the geospatial effect, and extracted the Watanabe-Akaike information criterion (WAIC) as a model adequacy metric.

#### Global multi-disease models

We first fitted general models to infer drivers of risk for all disease outbreaks (n = 49,239 after excluding early and spatially-imprecise records), with 50,000 background points across the global study area, not differentiating between specific diseases. To examine the potential confounding effects of local detection processes and broad-scale patterns of reporting effort on inferred drivers, we fitted three submodels: (1) including only socio-environmental covariates, i.e. with no outbreak detection-specific covariates (urban cover and healthcare travel time) and no geospatial effect; (2) adding a geospatial effect but no local outbreak detection-specific covariates; and (3) a full model with outbreak detection covariates and a geospatial effect (Figure 2). For comparison and sensitivity checking, we also fitted the full geospatial and detection covariate model for subsets of pathogens defined as either zoonotic (principally transmit to humans from an animal reservoir; n = 26) or vector-borne (transmitted to humans by arthropod vectors irrespective of host, i.e. also including anthroponotic arboviruses such as dengue fever; n = 20), whose risk is expected to be tightly coupled to local ecosystem characteristics (Extended Data Figure 3).

#### Individual disease-specific models

We fitted individual models for all diseases except Hendra virus disease, for which the number of human outbreak points was too low for reliable model fitting (Extended Data Table 1). The process of inference of drivers for each individual disease (n = 31) was as follows. First, we fitted separate geospatial models which included each covariate individually (“univariable”) plus a geospatial random effect to account for the broad geographical pattern in outbreak occurrence, but not possible finer-scale confounding by other variables (particularly detection proxies). We then fitted two hypothesis-driven multivariable geospatial models including either the broad (“majority rule”) or stricter (“top-ranked”) drivers from the coauthor exercise (Extended Data Figure 4). Because of the strong *a priori* expectation of detection bias driven by health systems proximity and accessibility, all hypothesis-driven models included both travel time to healthcare and urban cover; except in instances where these were highly collinear with each other; in these cases, the driver identified as most important in the hypothesis-generation exercise was selected. For all models where forest loss, cropland expansion or urban expansion were hypothesized as drivers, we also respectively included either forest cover, cropland cover or urban cover to account for the inherently spatially correlated process of land use change. For most diseases, urban expansion and urban cover were highly collinear at the scale of this analysis (Pearson’s *p* >0.8) so urban expansion was almost always excluded from multivariable models. For diseases with strongly hypothesized associations to specific livestock, the livestock covariate was based on gridded data for only the most relevant livestock type(s) (e.g. poultry for influenza, ruminants for Rift Valley fever; Supp. Table 1, Methods); the exception was MERS, as gridded camel density data are not openly available. Across all diseases, both the multivariable and hypothesis-driven models always reduced WAIC relative to a model including only a geospatial effect (i.e. including covariates improved model fit).

### Examining compound drivers across diseases

For many infectious diseases, synergistic interactions between drivers may be necessary to align the conditions for spillover and emergence risks (for example, high livestock densities in fragmented forest landscapes for bat-borne henipaviruses^7^). Improving geospatial prediction for emergence risks requires accounting for how compound drivers align to create local foci of pathogen transmission. To examine this question we visualized patterns of co-occurrence between drivers across all 31 individual modeled diseases. We generated a unipartite network with drivers represented as nodes, and with edges between driver pairs weighted by the number of diseases for which each pair of drivers co-occurred (i.e. when both drivers had 95% credible intervals not overlapping zero), for all diseases (Figure 4a) and for subsets of either directly-transmitted zoonoses (Figure 4b) or vector-borne zoonoses (Figure 4c). In parallel, to examine observed autocorrelation among putative drivers at global and regional scales, we generated a matrix of pairwise Pearson correlation coefficients between each pair of scaled covariates across 50,000 background points globally, or subsets of background points within five regions containing most of our data (North America, Latin America and the Caribbean, sub-Saharan Africa, East Asia and Pacific, and South Asia). These were used to visualize unipartite networks of pairwise driver correlations, with edges weighted by Pearson coefficient magnitude (Extended Data Figure 8).

### Limitations of data and methodology

Because the goal of the study was to apply a general, standardized analysis framework across a variety of diseases with very different quantities and types of data, we encountered several important but irreconcilable methodological constraints that are significant to interpretation of our results, as well as to inference of spatial drivers of disease emergence more broadly. Firstly, the datasets for many diseases (especially rare and high-consequence pathogens) are very small and spatially biased towards surveillance hotspots. We adjusted for these biases using geospatial random effects and proxies for detection processes, but these are imperfect descriptors for complex processes, and some residual confounding might remain unaccounted for (e.g. health systems access is influenced locally by many factors other than proximity, and clinical index of suspicion and accessibility of diagnostics is often highly geographically variable for many rarer, non-specific febrile illnesses). Relatedly, it was often not possible to combine multiple data sources for the same disease without creating a geographical imbalance in the distribution of points, so our analyses were mostly restricted to datasets that were usually broad in scale but lacked granular information about transmission intensity (e.g. most georeferenced outbreak datasets) or sometimes locally comprehensive at the expense of geographical breadth (e.g. hantavirus cardiopulmonary syndrome [HCPS], which was restricted to Brazil and Argentina due to available surveillance data, despite hantavirus infections occurring worldwide^103^). Notable exceptions where we were able to combine point and polygon data from more than one source without substantial issues included Lassa fever, Crimean-Congo hemorrhagic fever, HCPS, Chagas disease (acute), and yellow fever (Extended Data Table 1, Supp. Table 1).

Importantly, some of the most relevant variables thought to shape risk for many high-consequence epidemic zoonoses either have not (hunting pressure outside the tropics) or cannot (wildlife trade) be readily translated into global geospatial covariates that accurately reflect their relationship to infection risk. For example, the impacts of wildlife trade and markets on disease risks can be spatially diffuse and transboundary, involving multiple actors at multiple points along commodity chains from capture to sale^104^; consequently, quantifying how these activities shape the spatiotemporal dynamics of zoonotic spillover may require substantially different analytic approaches than what is possible with this study’s geolocated outbreak event data. (However, we also refer to other work that has highlighted instances where wildlife trade has been overstated as a driver of spillover risk.^20^) Similarly, coarse modeled spatial proxies for hunting pressure such as the tropical defaunation index we used in this study^60^ probably more closely reflect commercial rather than subsistence hunting activities, even though the latter may often be more important in driving zoonotic spillover (e.g. rodent hunting and exposure to Lassa fever and mpox); our study’s sparse and ambiguous results for tropical hunting pressure (Extended Data Figure 5; Supp. Figure 1) should be interpreted with this limitation in mind.

Developing a common analysis framework also led to the loss of information from some datasets, through reducing case surveillance data (with number of cases) to a binary annual outbreak indicator, (i.e. losing potentially valuable information on transmission intensity). Although necessary for a standardized framework, this could feasibly erode the reliability and accuracy of inference. We therefore conducted a model comparison test, examining how reducing the data’s information content affects inferred drivers for a set of relatively well-reported diseases (4 arboviruses in the USA using CDC ArboNET data). For each disease we compared coefficient estimates between full geospatial models of county-level case incidence, and our outbreak event risk modeling framework. County-level total case incidence across the surveillance period (2000-2020) was modelled using a negative binomial (West Nile fever) or zero-inflated negative binomial likelihood (LaCrosse encephalitis, Powassan encephalitis and Jamestown Canyon encephalitis), including an offset of log population, and a fitted geospatial random effect to account for unexplained geographical variation, again implemented using INLA. We found that most significant socio-environmental effects from a full case incidence model (i.e. reflecting transmission intensity) remained detectable even in a dataset reduced to binary outbreak occurrences with background locations (Extended Data Figure 7). This test improved our confidence that our modeling approach is sufficient to capture key spatial drivers of risk, despite this information loss.

Nonetheless, given data sparsity for many infections, it was not possible in this standardised framework to account for temporal dimensions of causality (e.g. time-specific climate or land change effects), such as by aligning covariate and case data in time; and/or adjusting for temporal patterns of detection through spatiotemporal random effects. This kind of analysis is feasible and fruitful when modeling case surveillance time series for better-surveyed infections (including some in our study such as *Borrelia burgdorferi* or West Nile fever), but it was not possible to apply this consistently across diseases, given the extreme sparsity of outbreak data for infections like Ebola, Marburg, Hendra, and Nipah virus disease. Rather than solely a limitation of this study, this is a more general problem for attribution of outbreak drivers for rarely documented but high-consequence infections, that currently hinders our capacity to, for example, robustly link recent deforestation to viral zoonosis outbreaks. Improving both fundamental eco-epidemiological research, and strengthening healthcare access, diagnostics and surveillance in underserved areas, will be needed to fill these gaps.

## Code and Data Availability

All code, data (where not subject to sharing constraints) and disease-specific results objects (e.g. CSVs of parameter estimates; rasters of fitted geospatial effects) are available on GitHub at github.com/viralemergence/fingerprint-preprint.

## Author contributions

Conceptualization: RG, SJR, GFA and CJC.

Study design and methodology: RG, SJR, CJC, DP, RLM, MPF, CHT, and BAH.

Hypothesis exercise design: SJR, CJC, and CAL.

Hypothesis exercise participation: RG, SJR, DP, MPF, RLM, GFA, DJB, HC-E, MC, EAE, HKF, BAH, ENH, KEJ, RK, AK, DL, CAL, JL, JPM, DWR, DR-A, BVS, SNS, and CJC.

Disease data processing: RG.

Modeling and analysis: RG.

Visualization: RG and CJC.

Data contribution: RG, DP, MPF, RLM, BVS, ENH, JPM, AS, EON, JKB, MC, JFM, DL, JL, DRA, AK, and CJC.

Writing (initial draft): RG, SJR, and CJC.

Writing (review and editing): all coauthors.

## Acknowledgements

This work was supported by an NSF Biology Integration Institute grant (NSF DBI 2021909 and 2213854), which supported RG, SJR, RM, GFA, DJB, EAE, HKF, BAH, SNS, and CJC, as well as the Verena Institute collaborative platform under which this work was organized (viralemergence.org). Further support came from the Trinity Challenge (RG, KEJ, DWR), the Wolfson Foundation (via a UCL Excellence Fellowship; RG), the Bill and Melinda Gates foundation (grant OPP#1181128; DMP), Bryce Carmine and Anne Carmine (née Percival) through the Massey University Foundation (RLM), the Wellcome Trust (award no. 101103/Z/13/Z; DL), and Schmidt Sciences (CHT). The funders had no role in study design, data collection and analysis, decision to publish, or preparation of manuscript. We thank Freya Shearer for discussions and substantial contributions to the data used in this study. We also thank the US CDC’s ArboNET platform for providing the data on US arboviruses.

## Extended Data

**Extended Data Table 1:**
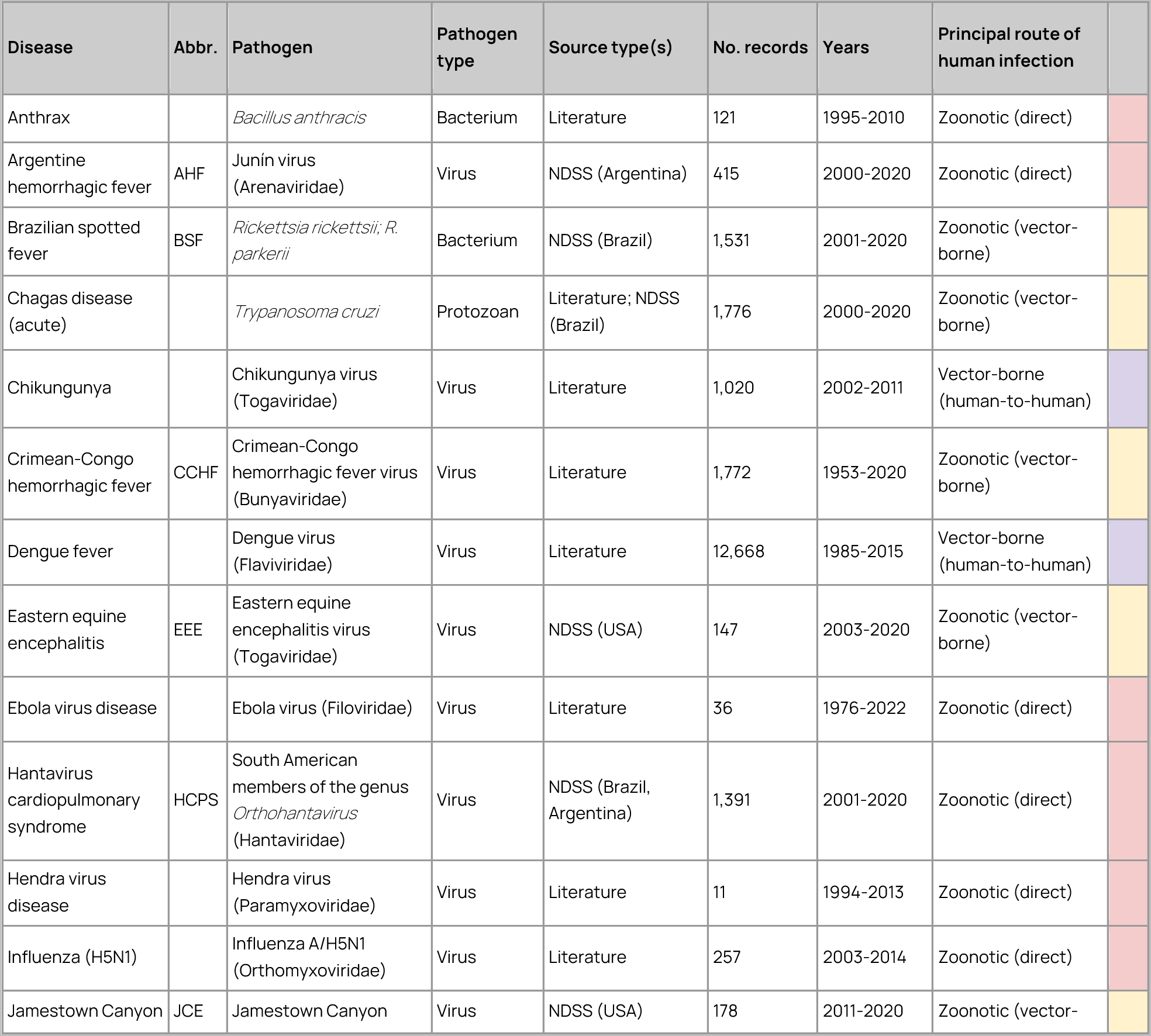
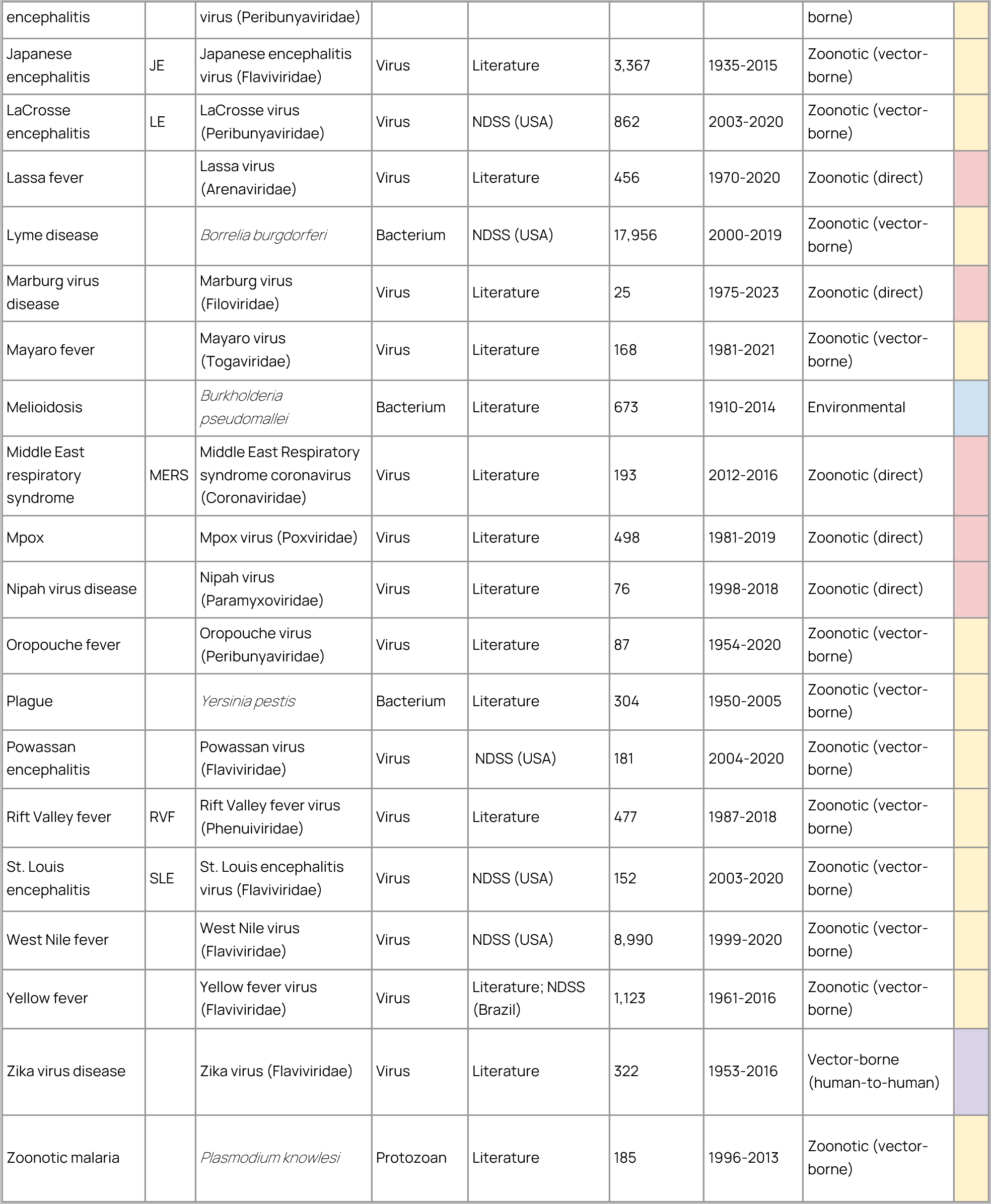
Database of outbreak events for 32 emerging infectious diseases. The table lists all diseases for which we were able to compile georeferenced human case or outbreak data, including the disease, pathogen(s), predominant transmission route to humans, number of outbreak event records and time period. A fuller set of data source descriptions with information about open accessibility for each source dataset is provided in Supp. Table 1. Most disease data were from a single source, but for several diseases we were able to combine and harmonize data from across more than one source database, as shown in the table (Extended Data Figure 1; e.g. hantaviruses; Lassa fever).This table includes all data points across all years (including prior to 1985) and regardless of spatial precision; prior to modeling, these data were subsequently subset to post-1985 records to better harmonize with covariate layers, and records with very low spatial precision were excluded (Methods). Abbreviations: NDSS - national disease surveillance systems.

**Extended Data Table 2:**
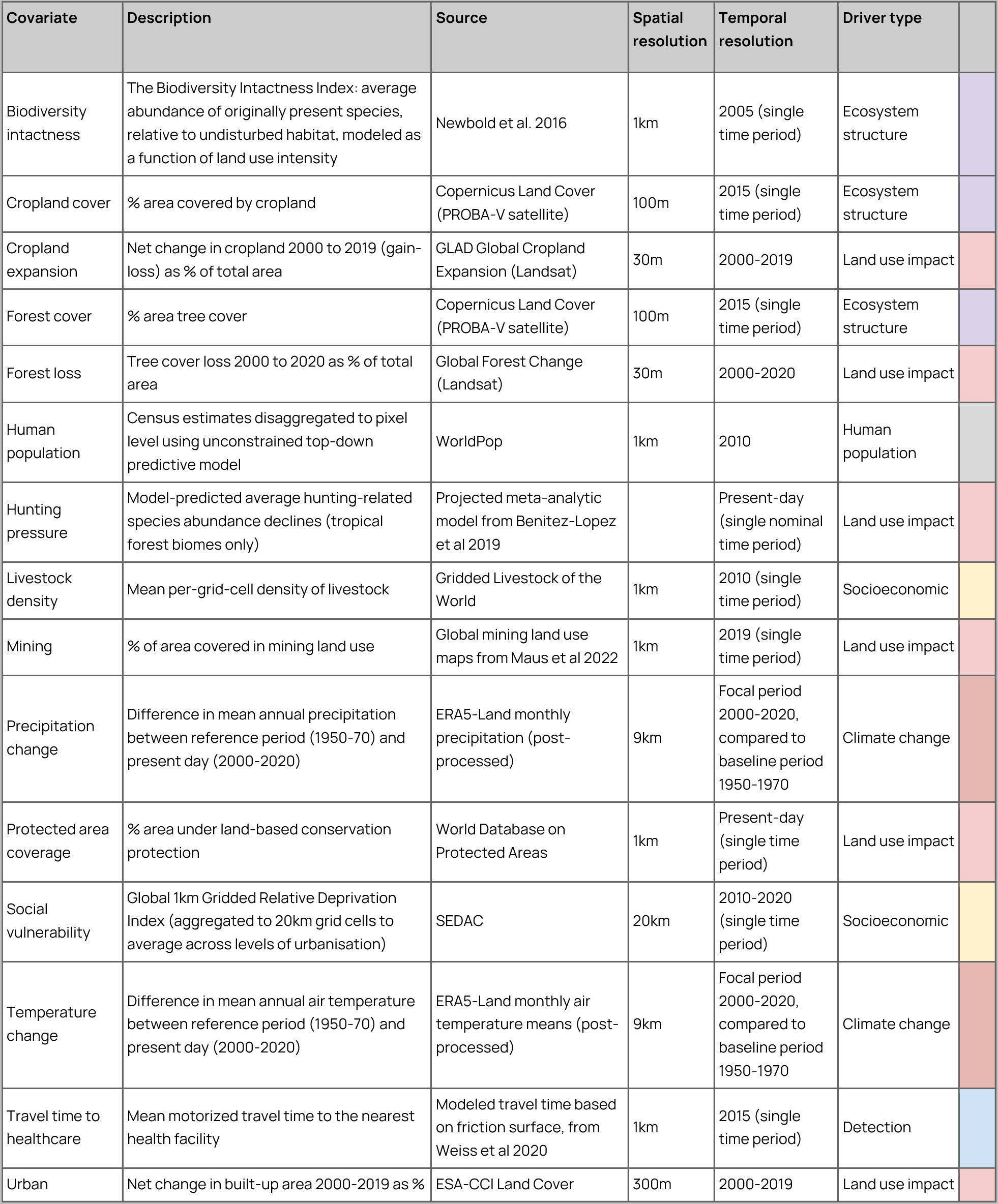
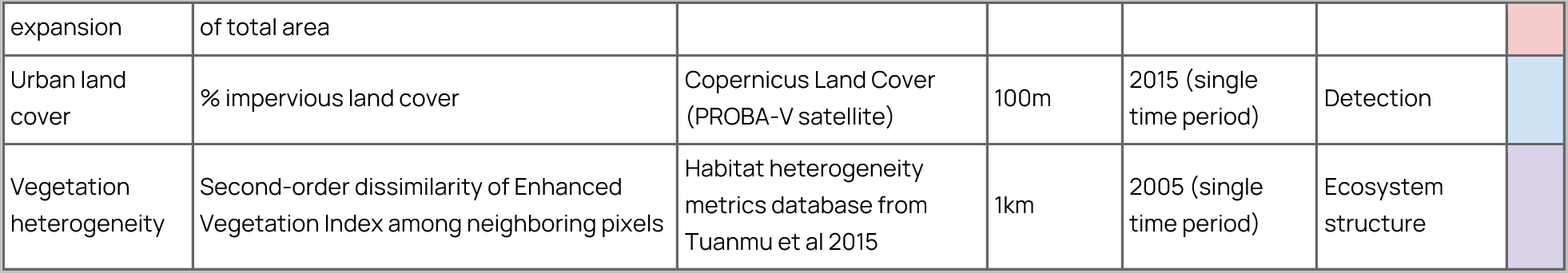
Socio-environmental covariate data sources. Table provides descriptions of the socio-environmental covariates used for each hypothesized driver, including the source, precise description, and spatial and temporal resolution. Information on open accessibility for each covariate is provided in Supp. Table 3.

**Extended Data Figure 1:**
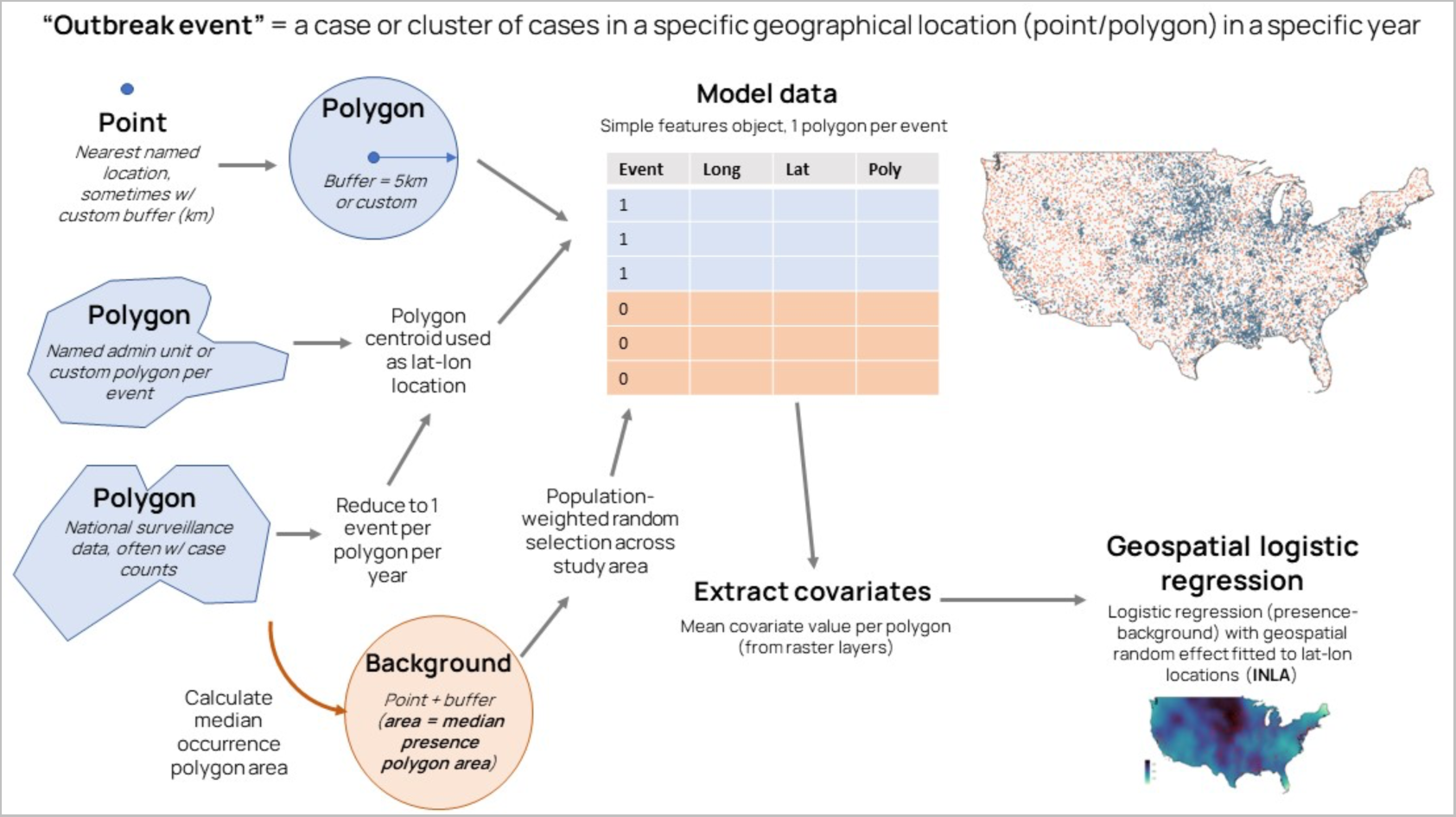
Bringing diverse disease case and outbreak data sources into a common analytical framework. Disease data sources included georeferenced case or outbreak event locations in point format (nearest named location), case or outbreak event occurrences within named administrative polygons, and case surveillance data at administrative polygon levels from national surveillance systems (sources variously shown in blue). Each contains different information about transmission intensity and different levels of geographical precision, which necessitated bringing different data types into a common, standardized analytical framework, shown in this figure. All outbreak locations (whether natively point or polygon) were converted into polygons (blue) and any polygons covering too large a spatial area were excluded as too imprecise (typically > 5000 km^2^, but up to 20,000 km^2^ for some data-deficient diseases as a compromise to retain as much data as possible; Methods). Background points were generated across the study area weighted by population (Methods, Extended Data Figure 2, map shown is for West Nile fever), then buffers were created around background locations to cover the same median area as the presence locations (orange), to ensure covariates were averaged across a comparable spatial area for both occurrence points and polygons and background locations. For each polygon the mean value of each raster covariate was calculated across the entire polygon, and used as input to geospatial logistic regression models.

**Extended Data Figure 2:**
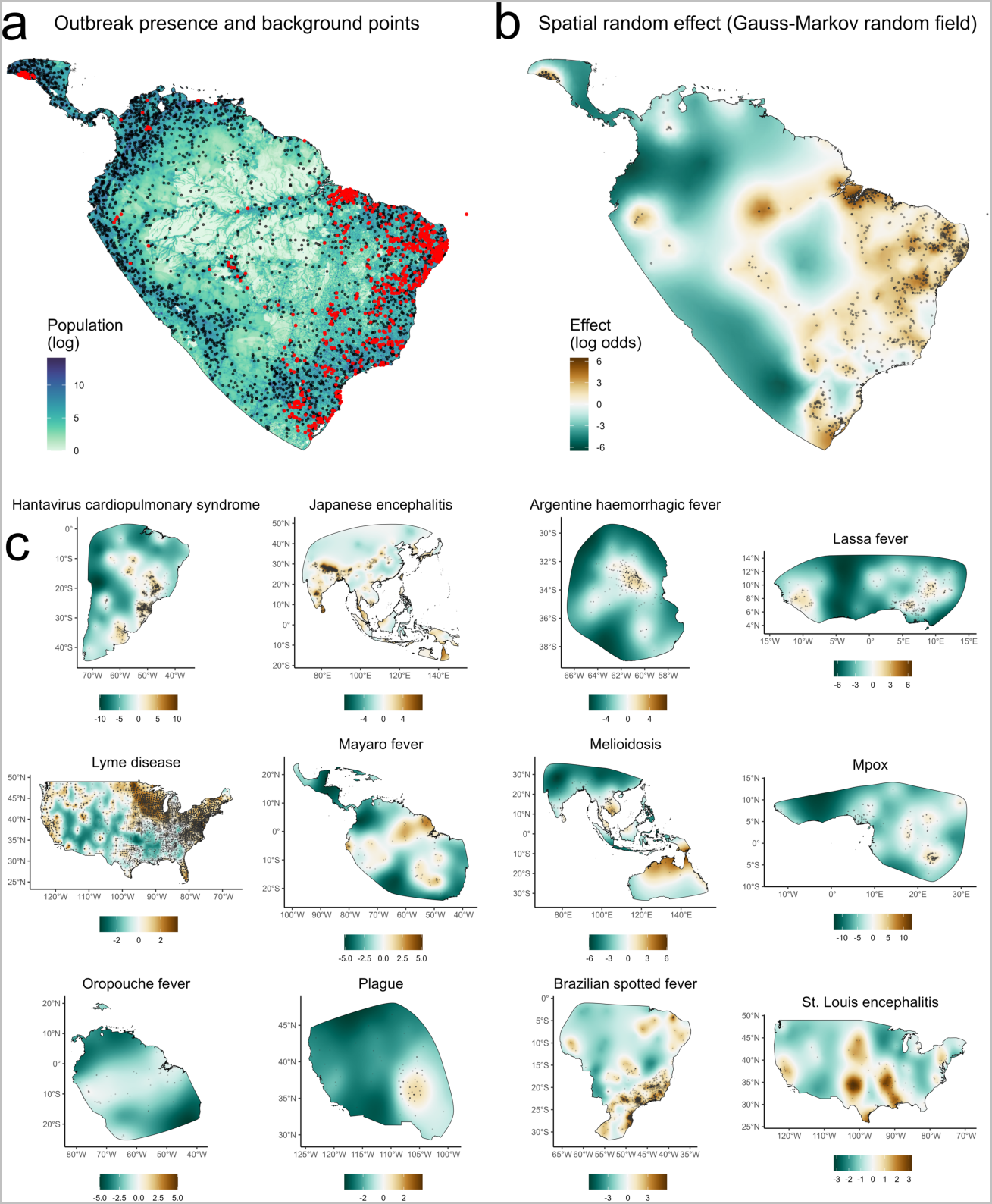
Case-control and geospatial model design for a subset of diseases. Geospatial logistic regression models were fitted to estimate the effect of covariates on the log odds of outbreak event occurrence (red points). The top row shows an example of model design for acute Chagas disease in Central and South America. Since outbreaks are presence-only data, we generated background points through randomly sampling 1 km grid cell locations across the study area (black border) weighted by log human population (left panel; shown as black points) to create a pseudo case-control design (i.e. comparing socio-environmental conditions at outbreak locations to the background distribution of conditions experienced by human populations overall) (A). To account for unmeasured factors shaping broad-scale outbreak geographies, models included a continuous geospatial random effect (Gauss-Markov random field; fitted field for Chagas disease is shown in top right panel) (B). Additional subpanels show fitted geospatial effects from the hypothesis-driven (“top ranked”) models for 12 randomly-selected diseases (C). Shading denotes the marginal contribution to outbreak risk (log odds scale), with brown denoting higher risk, and green denoting lower risk. Observed outbreak event locations are overlaid as gray points.

**Extended Data Figure 3:**
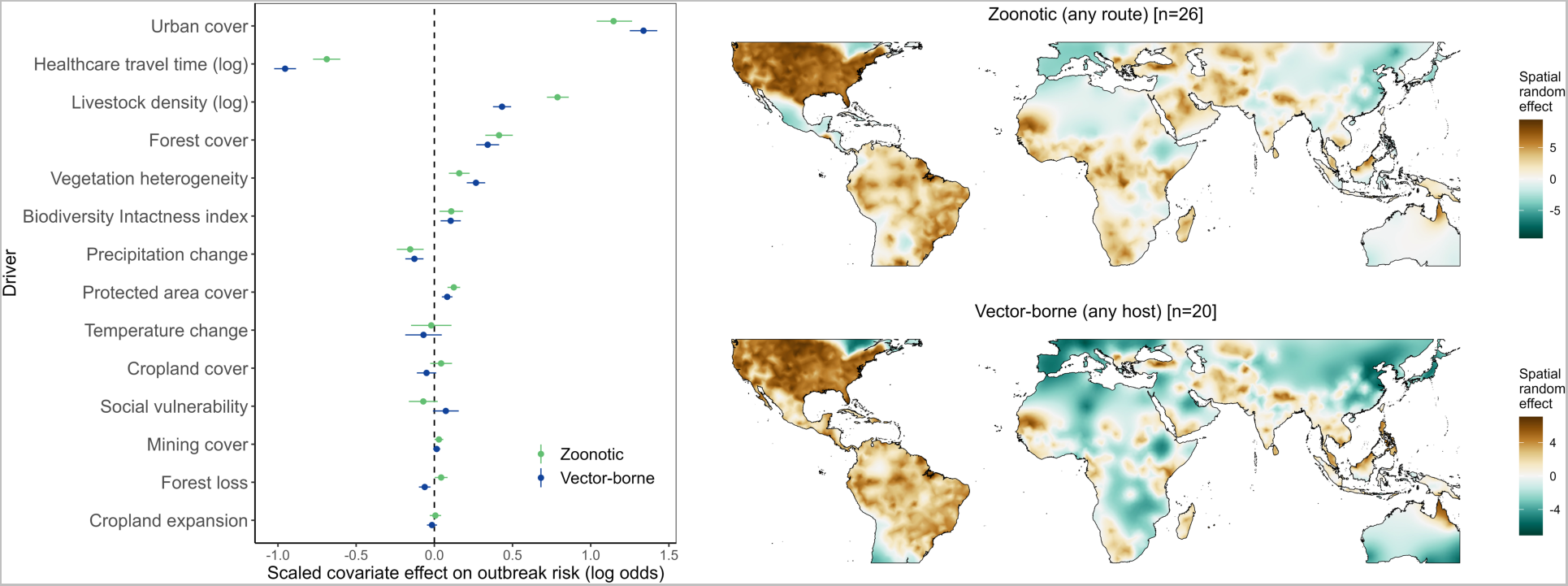
Global drivers of emerging disease outbreaks across different transmission groups. Replicating the analysis of Figure 2 (main text), global geospatial models were fitted separately for groups of diseases defined non-exclusively as either zoonotic (non-human animal reservoir with any mode of transmission; n = 26 diseases, 36,577 outbreak points) or vector-borne (transmitted by invertebrate vectors regardless of host, i.e. including principally anthroponotic arboviruses such as dengue; n = 20 diseases, 45,556 points). Points and error bars show linear fixed effects of scaled covariates (posterior marginal mean and 95% credible interval) from Bayesian logistic regression models fitted to all outbreak points, with point color denoting transmission group (zoonotic or vector-borne). Slope estimates denote the effect of each scaled covariate on spatial outbreak risk. Fitted geospatial random effects for each model (Gauss-Markov random field) are visualized as maps (color scale denotes marginal contribution to outbreak risk on the log-odds scale).

**Extended Data Figure 4:**
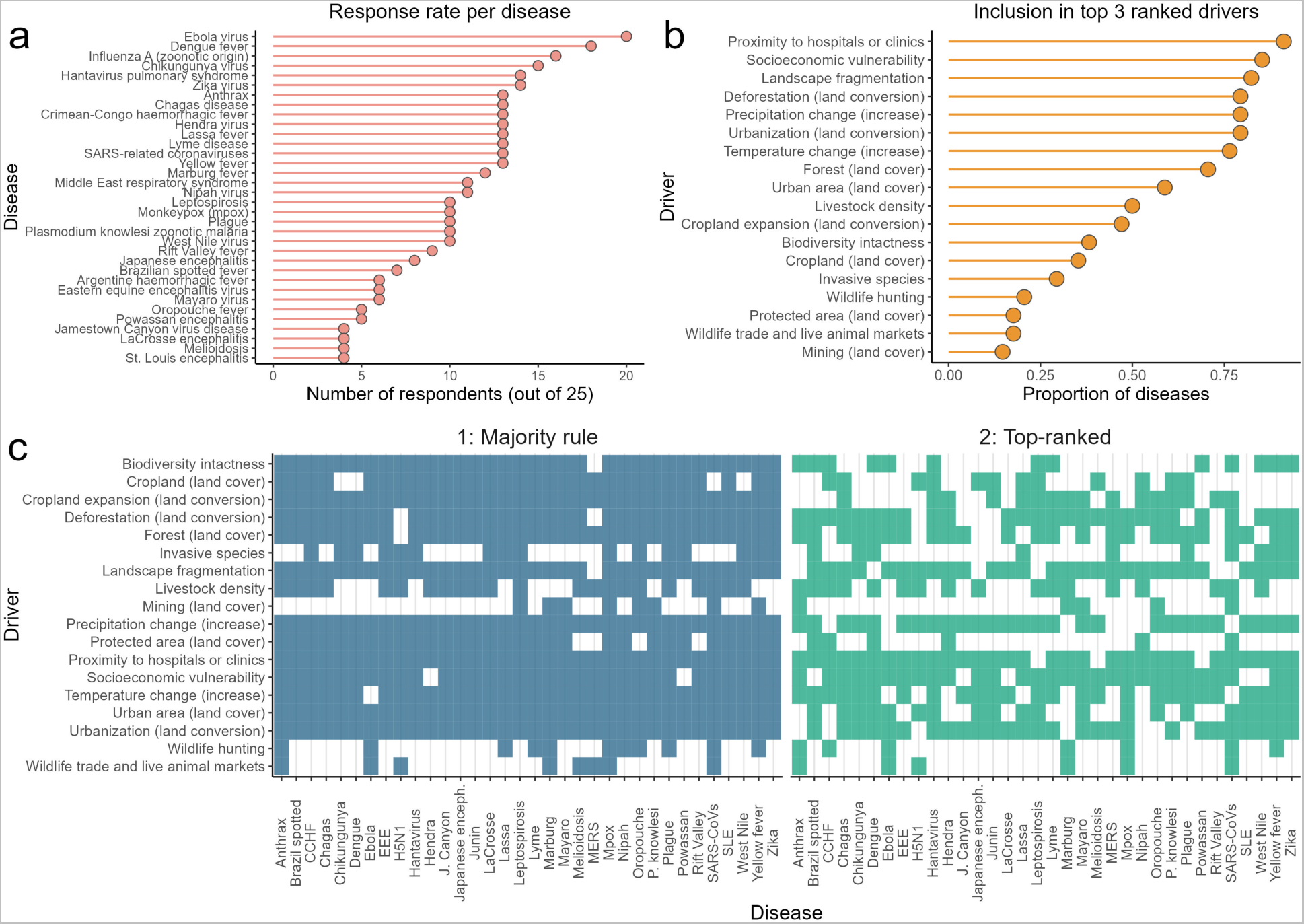
Hypothesised socio-environmental drivers for emerging infectious diseases from a collective hypothesis-generation exercise. To ensure our analyses tested appropriate, ecologically-plausible drivers for each disease, we used a structured form-based hypothesis exercise completed by the majority of coauthors (n = 25 out of 31; Methods). Respondents had the option to either fill in the form or leave blank for each disease (diseases names provided were as in panel A). There was substantial variability in response rates (A), with most responses for better-studied or widespread diseases (e.g. Ebola, dengue, influenza A) and vice versa. Respondents ranked each driver effect as “positive”, “negative”, “none” or “don’t know” and additionally were asked to select the top 3 most important drivers for each disease. Health systems access and socioeconomic vulnerability were the most commonly top-ranked drivers, followed by fragmentation, deforestation, urbanization and climate change (B; shows the proportion of diseases for which each driver was ranked in the top 3 by at least 1 respondent). Bottom panels (C) show hypothesized drivers to test for each disease based on two schemes: a broad “majority rule” criterion (drivers for which more respondents stated any effect than no effect) and a stricter “top ranked” criterion (all drivers that were ranked among the top 3 by at least 1 respondent).

**Extended Data Figure 5:**
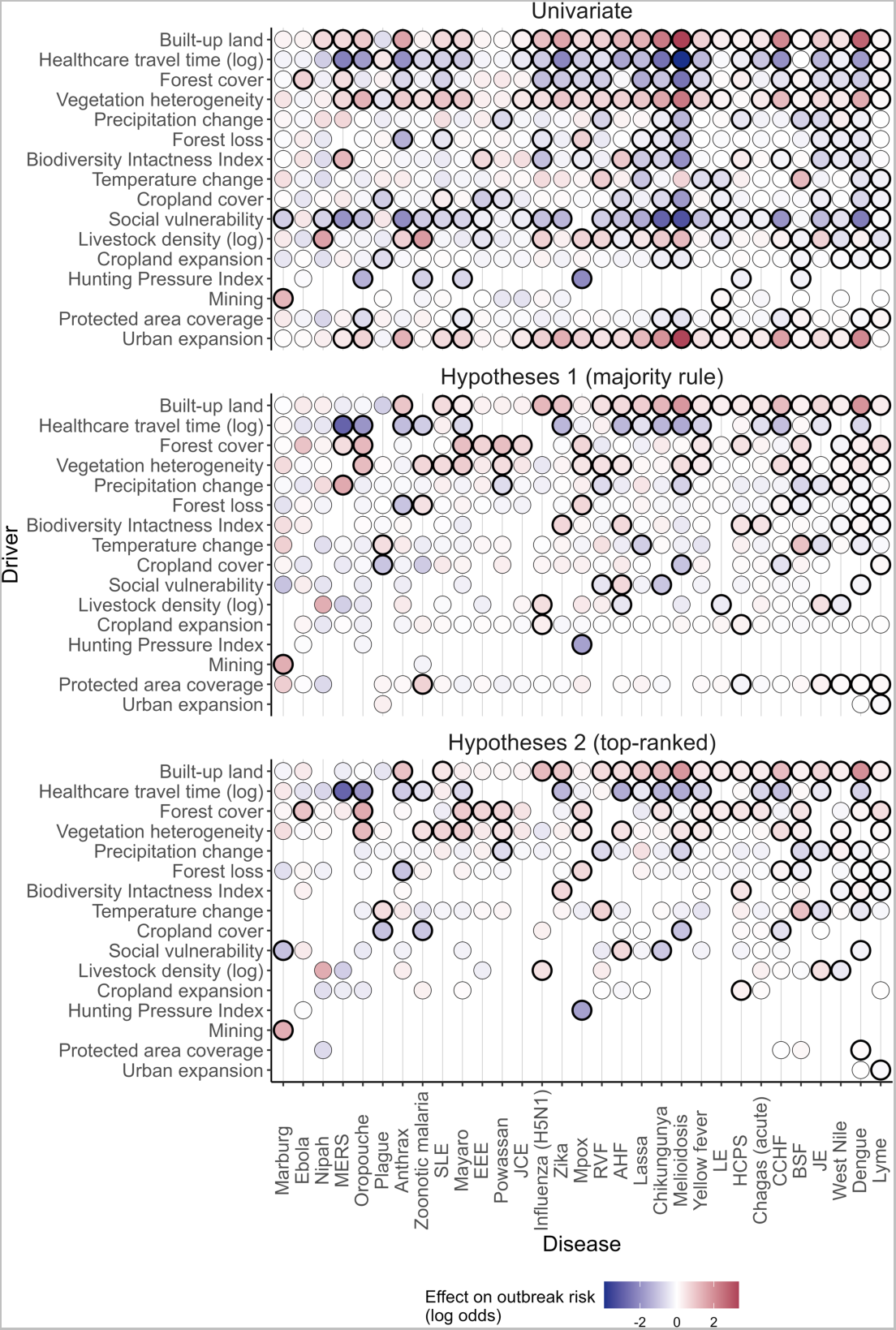
Estimated posterior mean effects of socio-environmental drivers of emerging infectious disease outbreaks, in univariable and hypothesis-driven models. Models were run in univariable driver-disease pairs (i.e. geospatial random effect plus each driver individually; top) and in multivariable models including two sets of hypothesized drivers identified through the hypothesis exercise (Extended Data Figure 4). These included a broader definition (“majority rule”: covariates that were identified by more respondents as having an effect on risk, than having no effect on risk; middle row), and a stricter definition (“top-ranked”: only covariates that were ranked among the top 3 drivers by any respondent; bottom row). Color represents the posterior mean linear effect of the scaled covariate (log odds scale), where red denotes increasing risk and blue denotes decreasing risk. Black borders denote evidence of a non-zero effect on risk (i.e. 95% credible interval not overlapping zero). Drivers are ranked by number of non-zero effects from the “top-ranked” models (top to bottom), and diseases are ordered from left to right by number of outbreak records (lowest to highest).

**Extended Data Figure 6:**
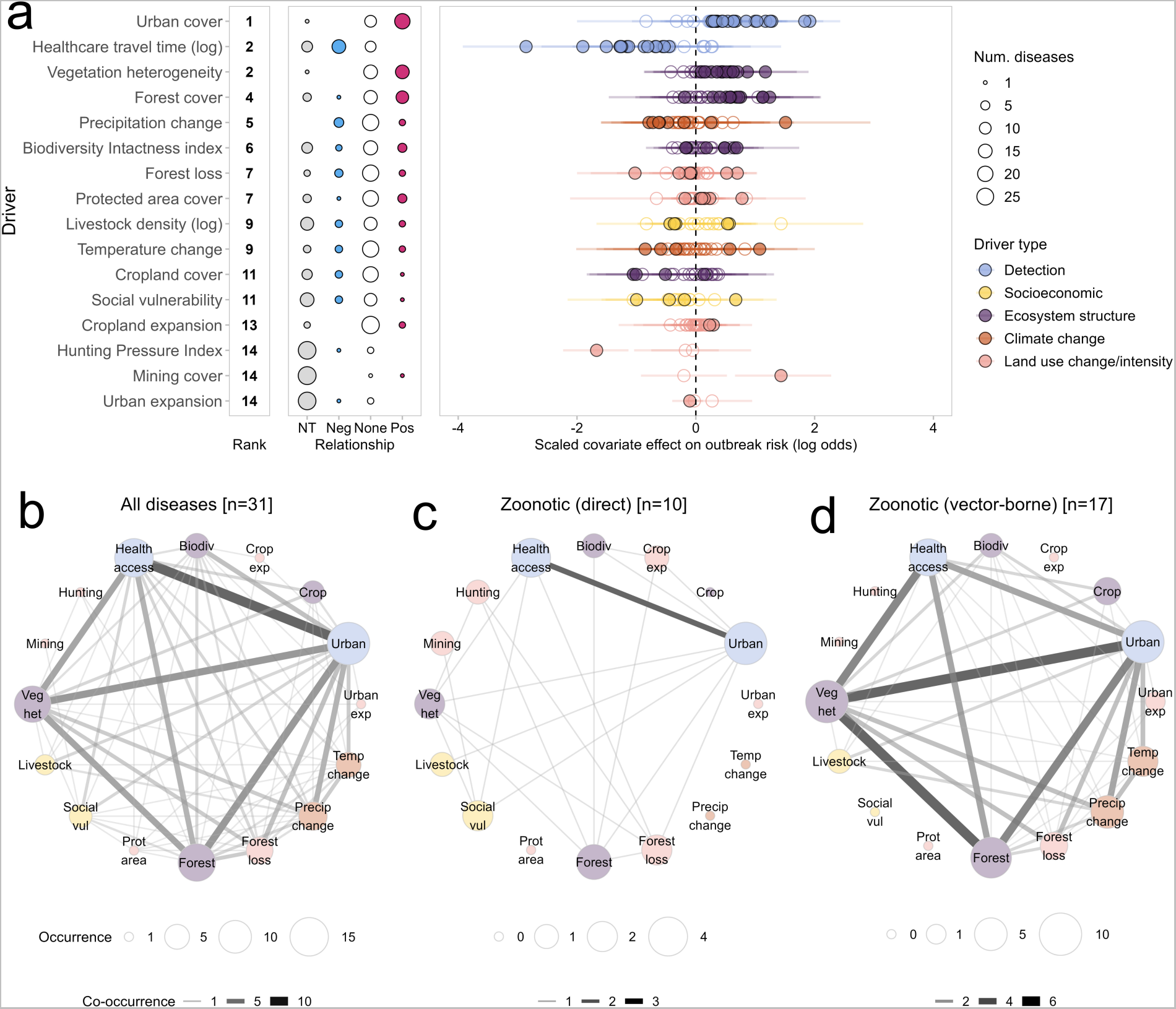
Drivers of outbreak risk for 31 emerging infectious diseases based on “top ranked” hypothesis criterion. The figure replicates the results from main text Figures 3 and 4, but based on hypotheses generated using the stricter “top ranked” criterion (Extended Data Figure 4c). Top row (A): panels show ranked drivers by number of diseases with strong evidence of a relationship (right column), prevalence and directionality of driver effects (middle column, with point size denoting number of diseases), and posterior marginal mean and 95% credible interval for all tested diseases (right column, filled points represent evidence of a non-zero effect). See Figure 3 legend for full description. Bottom row (B-D): unipartite networks show the pattern of co-occurring drivers for all diseases; directly-transmitted zoonoses; and vector-borne zoonoses. Nodes represent drivers with size proportional to the number of diseases with evidence of a non-zero effect; edge weight denotes the number of diseases for which driver pairs co-occur. See Figure 4 legend for full description.

**Extended Data Figure 7:**
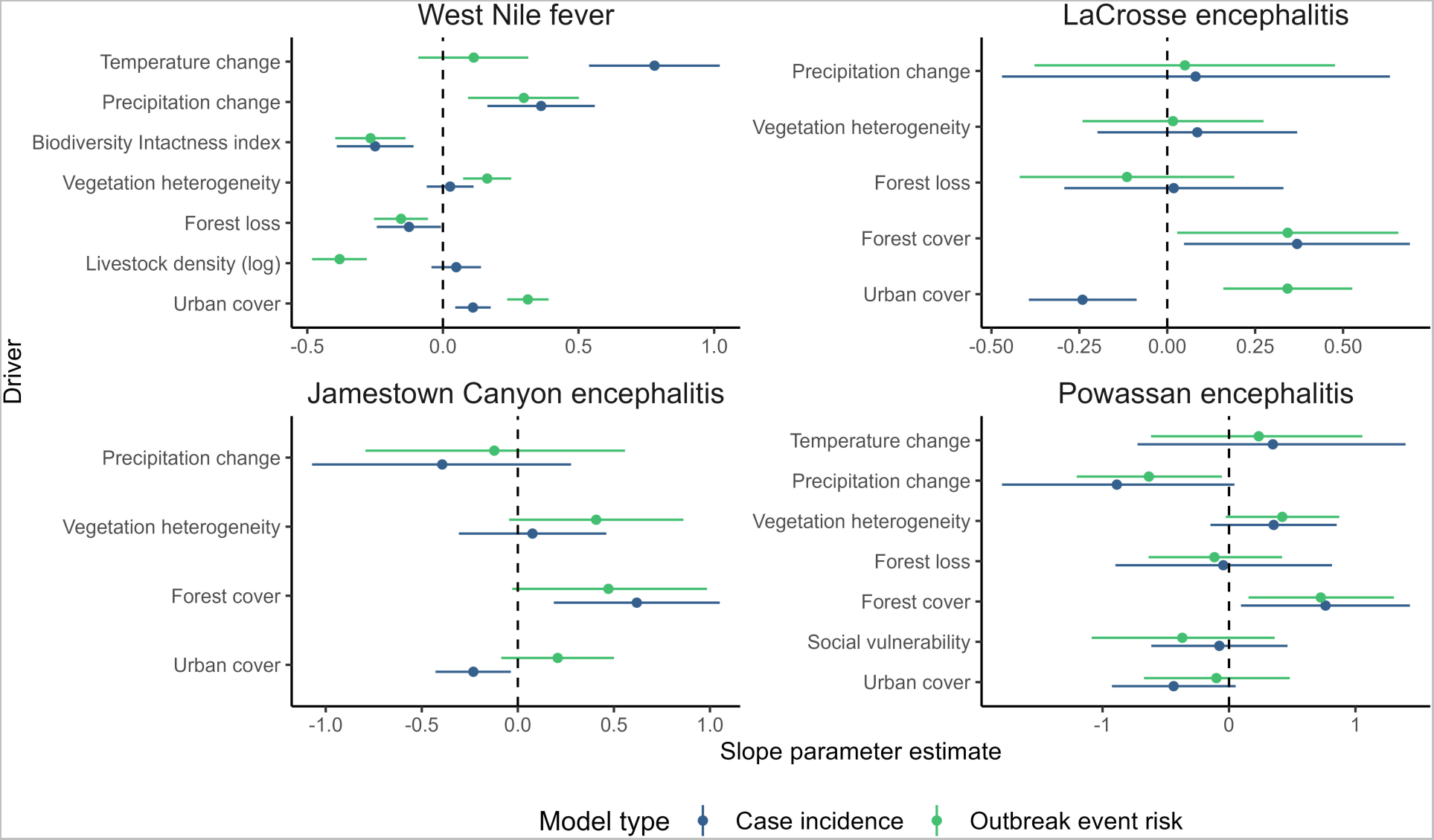
Comparison of detected socio-environmental drivers of disease incidence and outbreak event risk for arboviruses in the USA. Developing a common analytic framework based on outbreak events required discarding information about transmission intensity (i.e. number of cases) that is contained within national case surveillance datasets. To examine how this might affect inferred drivers, we compared coefficient estimates between full geospatial models of county-level case incidence, and our outbreak event risk modeling framework (Methods), for 4 diseases with varying quantities of case incidence data from the US CDC’s ArboNET surveillance platform. Incidence slope parameters (blue points and error segments) measure the inferred effects of each driver on observed log incidence (mean and 95% credible interval). These are shown alongside slope parameters from outbreak event models (green), which measure covariate effects on log odds of outbreak event occurrence compared to population-weighted background points (i.e. our standardized framework for this study; Methods). Drivers tested were based on the “top ranked” criterion in the hypothesis exercise (Extended Data Figure 4). Data: West Nile fever (annual 2004-2020; total cases=35,233; number of outbreak events=1,895; total counties included in model study area=3,084); LaCrosse encephalitis (annual 2003-2020; cases=1,369; outbreaks=306; counties=2,354); Jamestown Canyon encephalitis (annual 2000-2020; cases=225; outbreaks=112; counties=2,642); Powassan encephalitis (annual 2004-2020; cases=199; outbreaks=93; counties=1,146).

**Extended Data Figure 8:**
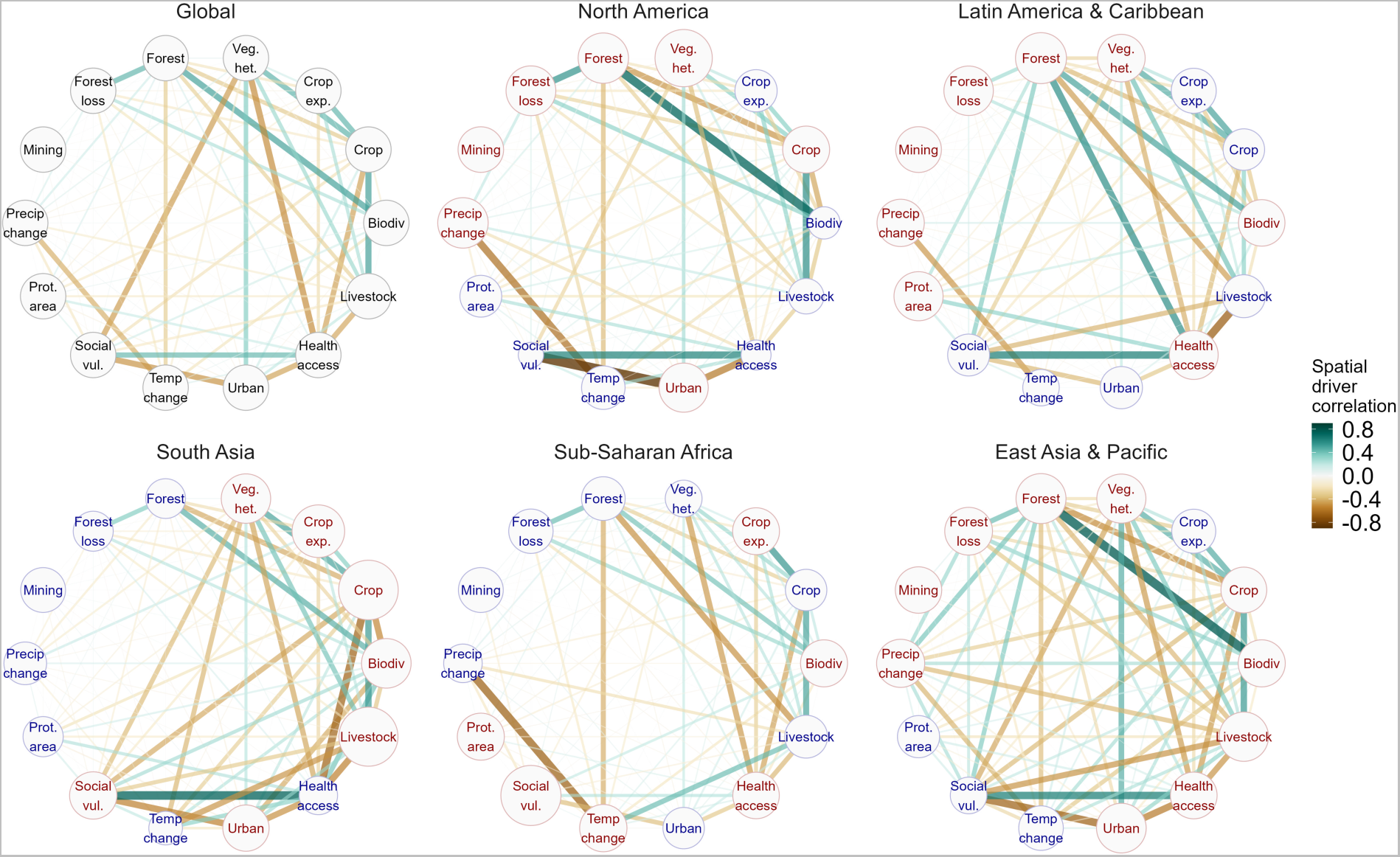
Clustering of emerging infectious disease drivers at global and regional scales. Networks show pairwise Pearson correlations between all driver covariates (nodes), with edge color showing direction and strength of correlation (positive in green, negative in brown) and edge weight denoting strength of correlation (i.e. absolute value). Correlations were calculated based on 50,000 population-weighted background points generated across the global study area (bounding box around all outbreak occurrences; Methods), with covariate values averaged across a 10km radius buffer around each point. Networks are shown using all background points (global) and separately for the five subregions containing most of the outbreak data. To visualize regional differences in covariate intensity per region, node sizes in region-specific networks are proportional to each covariate’s mean scaled value, with node text color denoting whether this was above (red) or below (blue) the global average (for example, North America has substantially lower mean social vulnerability than the global average across all points, and sub-Saharan Africa and South Asia substantially higher). Urban expansion was excluded as it was consistently highly correlated with urban cover (*ρ* > 0.85), and hunting was excluded as its restriction to tropical forest biomes resulted in a high proportion of missing values. Most variable pairs were uncorrelated or very weakly correlated (mean 13% of driver pairs with absolute *ρ* > 0.5, and 8% with absolute *ρ* > 0.7, across all regions).

## Supplementary Material

Supplementary Table 1: Disease data sources, links and access.

Supplementary Table 2: Hypothesis generation exercise form as completed by study coauthors.

Supplementary Table 3: Socio-environmental driver data sources, links and access.

Supplementary Figure 1: Forest plots of linear fixed effects from disease-specific multivariable models.

