## Supplementary Figures for "The anthropogenic fingerprint on emerging infectious diseases"

### Supplementary Figures for Gibb et al., “The anthropogenic fingerprint on emerging infectious diseases” (May 2024)

**Supp. Figure 1: Fitted linear fixed effects from hypothesis-driven models for all 31 modelled diseases.** Plots show inferred posterior marginal effects of scaled socio-environmental drivers on outbreak risk, with points and error bars showing posterior marginal mean and 95% credible interval. Results are from multivariable models including all hypothesized drivers under either a broader or stricter criterion derived from the hypothesis generation exercise (Methods, denoted by point colour). All models included a continuous geospatial random effect to account for unmeasured factors shaping spatial risk distribution (Methods).

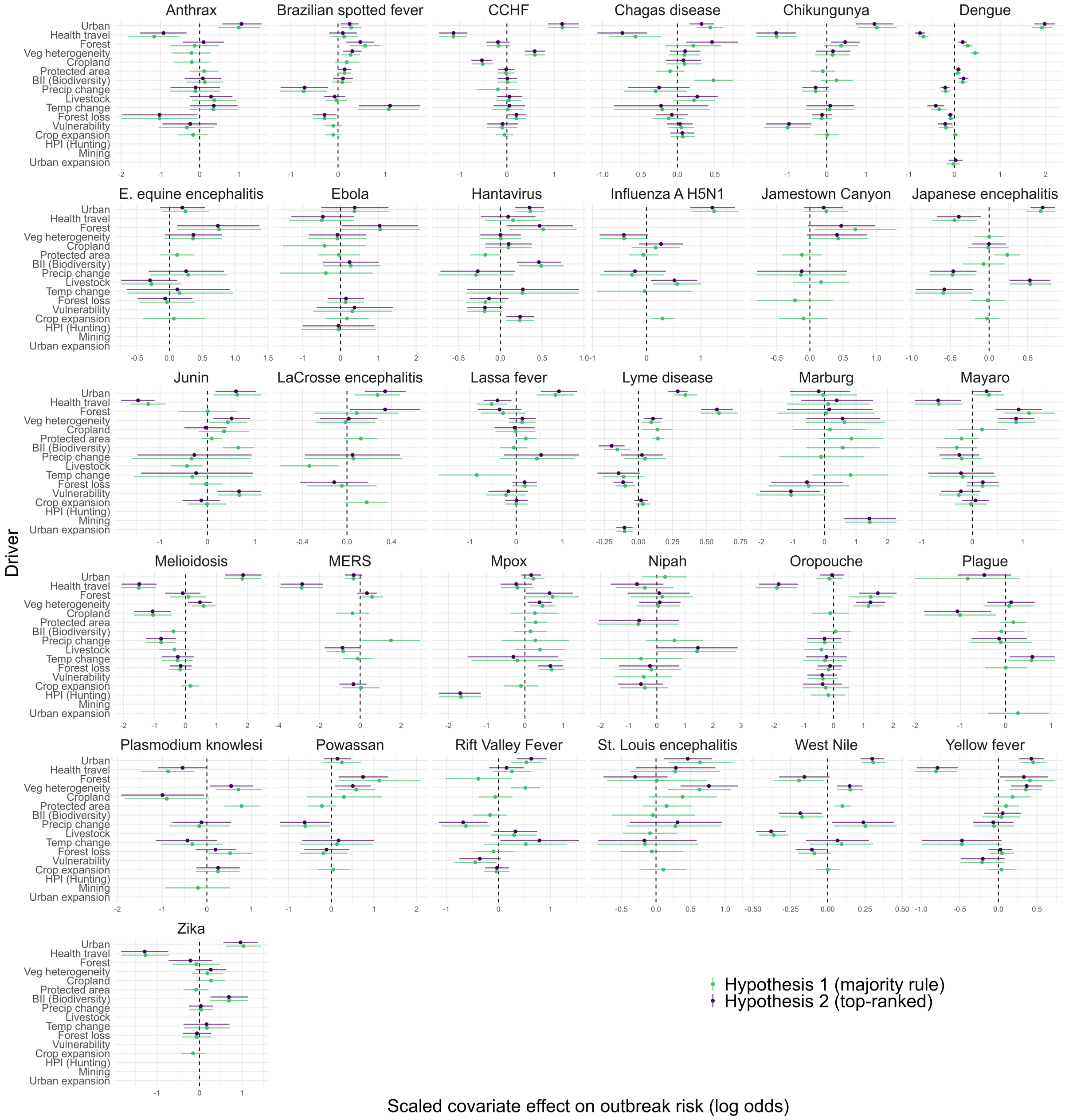
